# Changes in sensorimotor network dynamics in resting-state recordings in Parkinson’s Disease

**DOI:** 10.1101/2024.01.22.24301617

**Authors:** Oliver Kohl, Chetan Gohil, Nahid Zokaeia, Michele T. Hu, Anna C. Nobrea, Mark Woolrich, Andrew Quinn

## Abstract

Non-invasive recordings of magnetoencephalography (MEG) have been used for developing biomarkers for neural changes associated with Parkinson’s Disease (PD) but have yielded inconsistent findings. Here, we investigated whether analysing motor cortical activity within the context of large-scale brain network activity provides a more sensitive marker of changes in PD using MEG.

We extracted motor cortical beta power and beta bursts from resting-state MEG scans of individuals with PD (N=28) and well-matched healthy controls (N=36). To situate beta bursts in their brain network contexts, we used a time-delay embedded Hidden Markov Model (TDE-HMM) to extract brain network activity and investigated co-occurrence patterns between brain networks and beta bursts.

PD was associated with decreased beta power in motor cortex and decreased occurrences of the sensorimotor network, while motor cortical beta-burst dynamics were not changed. By comparing conventional burst and large-scale network occurrences, we observed that motor beta bursts occurred during both sensorimotor network and non-sensorimotor network activations. When using the large-scale network information provided by the TDE-HMM to focus on bursts that were active during sensorimotor network activations, significant decreases in burst dynamics could be observed in individuals with PD.

In conclusion, our findings suggest that decreased motor cortical beta power in PD is prominently associated with changes in sensorimotor network dynamics using MEG. Thus, investigating large-scale networks or considering the large-scale network context of motor cortical activations may be crucial for identifying alterations in the sensorimotor network that are prevalent in PD, and might help resolve contradicting findings in the literature.

**Highlights:**

- Sensorimotor network occurrences are decreased in Parkinson’s Disease.
- Motor cortical beta bursts occur during both sensorimotor network and non-sensorimotor network activations.
- Focusing on motor beta bursts occurring during sensorimotor network activations enables for better discrimination between controls and individuals with PD.
- The spatiotemporal details provided by large-scale network analysis may help to overcome discrepancies found in the PD literature.

## Introduction

Parkinson’s Disease (PD) is the fastest growing cause of neurological-related disability worldwide (Dorsey et al., 2018). Its pathological hallmark is the progressive loss of dopaminergic neurons and accumulation of Lewy bodies in the substantia nigra pars compacta compromising dopaminergic, regulatory processes of the basal ganglia (Hornykiewicz and Kish, 1987; Spillantini et al., 1997). This dysregulation has been linked to motor impairments such as Bradykinesia, Rigidity, Tremor, Postural Instability, and Dystonia (Hammond et al., 2007). A heterogenous spectrum of non-motor symptoms, e.g., hyposomnia, sleep disorders, depression, constipation, and cognitive impairments, can be observed in addition to the cardinal motor symptoms, as other brain systems are impacted (Aarsland et al., 2021; Schapira et al., 2017).

An important effort is that of defining clear, sensitive, and selective non-invasive biomarkers of PD pathology that can be measured in a wide range of individuals. Magnetoencephalography (MEG) is a promising candidate because it directly measures magnetic fields arising from cortical neural activity which may enable the assessment of synaptic integrity and functioning in local and large-scale networks (Lanskey et al., 2022). Such non-invasive biomarkers should prove useful for the early identification of disorder and treatment development and management.

Experimental investigations using invasive methods in individuals with PD and animal models have advanced our understanding of the neurophysiological underpinnings of PD pathology. Excessive low beta oscillations in the basal ganglia-thalamocortical loop that is underpinned by prolonged periods of dynamically occurring low beta synchronisation (=bursts) has been proposed to provide functional markers of PD (Hammond et al., 2007; Lofredi et al., 2023; Tinkhauser et al., 2017b). Observation of co-occurring STN and motor cortical bursts (Tinkhauser et al., 2018), paired with reports of exaggerated motor cortical beta power in animal models (Iskhakova et al., 2021; Sharott et al., 2005) and invasive motor cortical electrocorticogram (ECoG) recordings in individuals with PD (Crowell et al., 2012; O’Keeffe et al., 2020) pose the hypothesis that motor cortical beat power is exaggerated in individuals with PD.

Interestingly, global observations of motor cortical beta power made using non-invasive neurophysiological methods are inconsistent with the local observations in the invasive literature. For example, MEG and Electroencephalogram (EEG) studies that have investigated oscillatory changes in motor cortex have led to mixed findings. Some studies report no changes in motor cortical beta power (Shirahige et al., 2020), whereas others report decreased (Heinrichs-Graham et al., 2014; Vardy et al., 2011) or increased beta power (Hall et al., 2014; Pollok et al., 2012). Furthermore, the associations drawn between beta-power changes in motor cortex and motor symptom severity are also mixed (Boon et al., 2019; Geraedts et al., 2018). Other reports show that PD-related decreases in the motor cortical power-spectrum normalise following administration of dopaminergic medication (Cao et al., 2020; Heinrichs-Graham et al., 2014; Melgari, 2014).

Adopting a dynamic, burst-based approach, has not proven sufficient to harmonise findings. Analyses of motor beta bursts in non-invasive MEG recordings have generated inconsistent findings regarding patterns of changes in their dynamic properties: decreases in motor cortical beta-burst rates (Vinding et al., 2020), no group differences in burst rates but a steeper age-related decline in burst rates (Vinding et al., 2021), and increased burst durations and amplitudes in individuals with PD that normalised after deep brain stimulation have been reported (Pauls et al., 2022). Changes in burst rates were reported to scale with Bradykinesia symptom severity (Vinding et al., 2021, 2020), and burst amplitudes were associated with UPDRS hemi-body scores (Pauls et al., 2022).

This puzzle of inconsistent findings among the non-invasive studies, as well as between non-invasive and invasive studies, may be caused by many factors such as varying disease progression, symptomatology, assessment of intervention effects vs. HC-vs.-PD group contrasts, or scale of measurements. Especially given the large scale of M/EEG measurements, it is likely that activity in a single scalp region constitutes different types of activity, with each type associated with different brain networks.

We propose that the large-scale network context within which beta bursts occur is a fundamental factor to consider when investigating non-invasively measured motor cortical activity. Beta bursts recorded non-invasively from motor sensors likely include activity associated with brain networks beyond the sensorimotor areas. For example, previous MEG studies demonstrated beta power decreases that are linked to spectral slowing in central and posterior cortical areas in PD (Bosboom et al., 2006; Stoffers et al., 2007). Importantly, in conventional spectral and bursts analyses, a sensor or region of interest is pre-selected, meaning that the network context of beta activations is neglected. The network context, however, may be essential for dissecting relevant motor cortical activations and thereby pinpointing functionally relevant pathology. Considering the centrality and prevalence of motor symptoms in the diagnosis of PD, we hypothesize that more reliable markers of PD can be extracted by focusing on motor beta activations that occur specifically in the context of activity in the sensorimotor network.

In the present work, we investigate whether the large-scale network context of motor cortical activations, extracted with a time-delay embedded Hidden Markov Model (TDE-HMM) (Vidaurre et al., 2018b), helps to identify pathophysiological changes in PD. We test whether sensorimotor network dynamics are altered in individuals with PD. We further hypothesize that motor beta bursts may be associated with various large-scale network contexts, not all of which are closely linked to pathological changes in PD. By separating motor cortical activations associated with the sensorimotor network, it should be possible to tap into processes that are more closely related to the primary PD pathology.

We start by trying to replicate previous observed changes in static motor cortical oscillatory beta power and beta bursts extracted with amplitude thresholding. We then compare large-scale brain network dynamics extracted with a TDE-HMM between the Healthy Control (HCs) and PD group to test whether previously observed changes in burst dynamics can be observed in a sensorimotor large-scale network. Crucially, we finally investigate the co-occurrence patterns between large-scale networks and motor beta bursts to determine whether focussing on beta bursts associated with the sensorimotor network improves discrimination between individuals with PD and healthy controls.

## Materials and methods

MEG recordings from 8-minute resting-state scans with eyes-open of 28 participants with PD and 36 HCs, matched in age and education, were analysed for this study. The resting-state scans were collected alongside task-based data in two previous studies by our lab (Heideman et al., 2020; Zokaei et al., 2021). The scanning protocol for the resting-state scans of both studies was matched. MEG recordings were acquired at a sampling rate of 1000 Hz using a VectorView scanner. Individuals with PD were asked to withdraw from their dopaminergic medication from 7 p.m. at the night before the MEG-scan. Cognitive scores, measured with the Addenbrooke’s Cognitive Examination or Montreal Cognitive Assessment, did not differ significantly between the two groups. PD volunteers were recruited via neurological clinics in Oxfordshire (UK) and the Dementias and Neurodegenerative Disease Research Network (DeNDRoN). An overview of the exclusion criteria of both studies can be found in the SI Table 1. Both studies were approved by the NHS Oxford Research Ethics Committee and the Research Ethics Committee of the University of Oxford and followed the Declaration of Helsinki. All participants gave written informed consent before participating.

**Table 1.** Descriptive statistics of all participants included.

| Demographics | Healthy Controls | Parkinson's Disease |
| --- | --- | --- |
| N | 36 | 28 |
| Sex<br>(female/male) | 13/23 | 15/13 |
| Handedness<br>(right/left) | 34/2 | 25/3 |
| Age (years)<br>(mean, range) | 68.36 (60-80) | 68.5 (54-83) |
| Education (years)<br>(mean, range) | 15.36 (10-21) | 14.68 (10-24) |
| Years since Diagnosis (years)<br>(mean, range) | N/A | 2.86 (1-7) |
| UPDRS III motor score<br>(mean, range) | N/A | 30.36 (11-55) |
| Hoehn and Yahr scale<br>(mean, range) | N/A | 1.77 (1-3) |
| Levodopa-equivalent daily dose<br>(mg/day)<br>(mean, range) | N/A | 393.94 (150-900) |

N = number of participants; UPDRS = Unified Parkinson Disease Rating Scale

### Software

All steps of the preprocessing were performed in Matlab R2020a using the Matlab version of the OHBA Software Library (OSL, https://github.com/OHBA-analysis/osl-core), Statistical Parametric mapping (SPM12; https://www.fil.ion.ucl.ac.uk/spm/) (Friston et al., 2007), or the HMM-MAR toolbox (https://github.com/OHBA-analysis/HMM-MAR) (Vidaurre et al., 2016), TDE-HMMs were run with the HMM-MAR toolbox.

Analyses of the TDE-HMM outputs were performed in Python 3.10. HMM state descriptions were estimated using OSL-Dynamics (https://github.com/OHBA-analysis/osl-dynamics). GLMs contrasting the two groups, while controlling for confounds, were calculated with GLMTOOLs (https://pypi.org/project/glmtools/), and cluster-based permutation tests were calculated as implemented in MNE-Python (Gramfort, 2013). Analysis scripts used for the analyses will be shared on GitHub upon publication. A schematic of all the analyses performed in this paper can be found in Figure 1.

**Figure 1.**
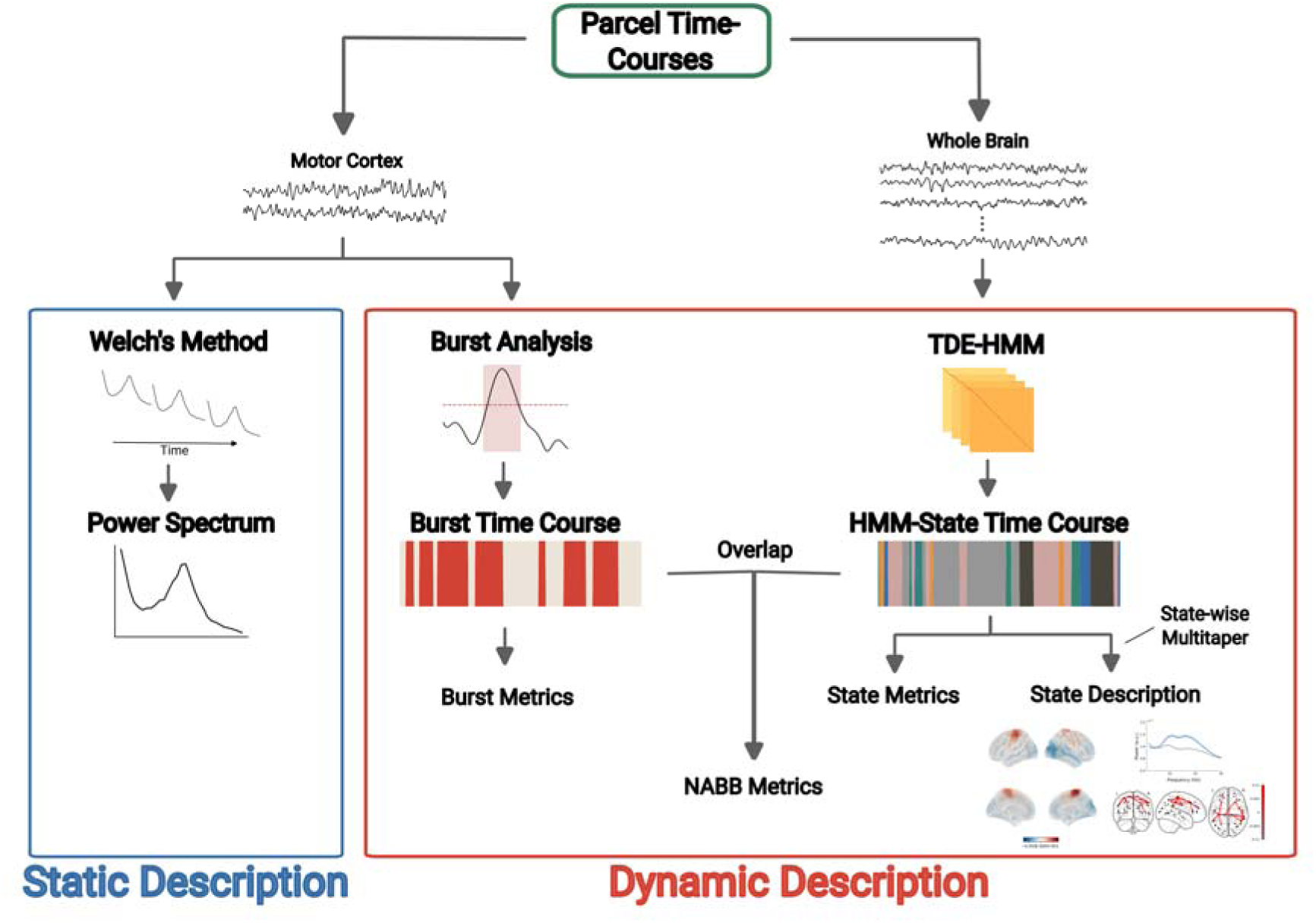
Schematic of the different analyses reported in this paper. All analysis were run on the parcellated and sign-flipped data. Welch’s Method was applied to time-courses of the left and right motor cortex to obtain a static description of motor cortical oscillations. The same time courses were used for the conventional amplitude thresholding bursts analysis to extract motor cortical beta-burst time courses. From these burst time courses, burst metrics describing dynamic features of these events were calculated. A TDE-HMM was applied to the time courses of all parcels to extract HMM-State time courses. Based on these state time courses, spectral state descriptions were calculated with a state-wise multitaper approach and state metrics were calculated to describe the dynamics of these states. Lastly, conventional beta bursts were sub-divided according to the HMM-State with which they co-occurred, and metrics describing the dynamics of these co-occurrences were calculated.

### Preprocessing

The MEG data were maxfiltered (Maxfilter 2.2) with the temporal extension option (--tsss) to attenuate components in the MEG signal originating from outside the head. The data were further transformed to a reference head position (--trans) and corrected for movements during the scan (--movecomp.) The maxfiltered data were converted into SPM12 format in Matlab for further processing. The MEG recordings were registered to the participants’ structural MRI scans with RHINO (Registration of Headshapes Including Nose). The data were downsampled to 400 Hz before applying a 0.1 to 125-Hz broadband filter and notch filters at 50 and 100 Hz using SPM12. Bad channels and bad segments were identified with osl_detect_artefacts. Across all participants, an average of 4 channels were rejected as bad channels and 5% of the recording duration was identified and removed as bad segments. To reject eye-movement and heartbeat artefacts in the data, OSL Independent Component Analysis (ICA) artefact removal was applied. ICA-Components highly correlated (>= .5) with the Electrooculogram or Electrocardiogram channels were rejected. In cases where no subcomponents were linked to the artefact channels or more than 4 channels were rejected, manual artefact detection was conducted on the ICA output. On average, 3 ICA components (SD = ±1) per participant were removed through this procedure. The MEG sensor data were then reconstructed without the ‘artefactual’ components and carried forward to the source analysis. These ICA-cleaned data have a reduced rank but are less affected by physiological noise sources.

Sensor normalization was applied to ensure equal contribution from MEGMAG and MEGPLANAR channels to the source localisation (Woolrich et al., 2011). A Linearly Constrained Minimum Variance (LCMV) vector beamformer (Van Veen and Buckley, 1988; Woolrich et al., 2011) with a Principal Component Analysis (PCA) regularisation rank of 50 projected MEG sensor data onto an 8-mm MNI152 standard brain template. Parcel-wise time courses were calculated for a weighted parcellation with 39 cortical regions (Colclough et al., 2015). For each parcel, voxel contributions were weighted according to respective parcel weights and the first principal component across voxels was specified as the parcel’s time course.

For the HMM analysis, the source reconstructed parcel time courses were broadband filtered to 1 to 45-Hz and downsampled to 250 Hz. We then performed symmetric multivariate leakage reduction, which corrects for both direct and inherited (so-called ghost interactions) (Palva et al., 2018) in spatial leakage (Colclough et al., 2015). Sign-flipping (Vidaurre et al., 2016) was applied next. Finally, the data were split into 10-second segments and generalized extreme Studentized deviate outlier detection (Rosner, 1983) was used to remove segments with exaggerated variance or kurtosis values. On average, 5% (SD = ± 3%) of the data were identified as bad segments. These preprocessing steps ensured that the time-delay embedded Hidden Markov Model analysis could accurately identify brain states without being influenced by outlier events. The resulting data were used for all subsequent analyses.

### Static Power

To explore the signal power across a range of frequencies, we used Welch’s method to compute power spectra. The algorithm was applied to each participant’s individually z-standardized parcel time course using a window length of 500 samples and a window overlap of 250 samples to obtain relative power per frequency bin. After averaging relative power spectral values across the two superior motor parcels for each participant, average power at each frequency bin was contrasted between the PD and control group with a t-test and cluster-based permutations (Gramfort, 2013).

### Burst Analysis

We applied a conventional amplitude-thresholding burst analysis to the two superior motor parcels to explore the dynamics of individual beta-power events underpinning spectral power in the time domain. For each participant, the preprocessed data were filtered between 18 and 25-Hz, based on the significant cluster obtained from the static motor cortical power group contrast. Next, we calculated amplitude envelopes of the time courses using the Hilbert transform and thresholded them at the 75th percentile, separately for each participant. Finally, bursts were identified as instances in which the threshold was crossed for longer than 1 cycle of the lowest frequency (18 Hz) in the frequency range of interest (Cole et al., 2019). This resulted in binary on-versus-off time courses, with 1 indicating samples during bursts and 0 indicating samples during burst-off periods. A bilateral motor cortical on-versus-off burst time course was created by assuming a burst occurrence at instances when a burst occurred in at least one of the motor parcels. The average burst overlap between motor cortical parcels was 69% (SD = ± 1%). Burst time courses of both motor cortical parcels were combined to enable for fair comparisons with TDE-HMM states, which were predominantly bilateral. Using the combined burst on-versus-off time course, we extracted the fractional occupancy (fraction of total time with bursts present), burst lifetimes (duration of burst occurrences), and state rates (number of burst occurrences per second). Additionally, we estimated motor cortical beta-power changes during bursts using a multitaper approach, calculating power weighted by burst on- and off- sets (Vidaurre et al., 2016). After subtracting the mean power across burst-on and -off periods, motor cortical beta-power estimates were averaged across both motor parcels to obtain beta-power changes during burst events.

### Time-Delay Embedded Hidden Markov Model (TDE-HMM)

#### TDE-HMM Description

We employed a time-delay embedded HMM to our data to investigate whether large-scale network information can be used to improve differentiation between the HC and PD group. TDE-HMMs extract dynamic brain states in a data-driven manner, with each state representing a large-scale functional network characterised by distinct multi-region oscillatory activity, i.e., distinct power covariations and phase synchrony patterns (Vidaurre et al., 2018b). Compared to conventional single-region beta-burst analyses, the HMM provides a richer description, by linking bursting activity in any region to the associated large-scale, multi-region network activity. In other words, each time a large-scale network occurs, it can be thought of as a transient bursting event of a large-scale, oscillatory network. The data-driven (unsupervised learning) nature of TDE-HMMs further obviates the necessity of setting *a priori* assumptions (i.e., region of interest, filter range, or beta-amplitude threshold) and thus reduces arbitrariness.

A detailed description of the inference process can be found elsewhere (Vidaurre et al., 2018b). In short, the standardised time series are described as a sequence of a defined number of hidden states, in which only one state is active at any moment. Each state is defined by an observation model that captures a distinct auto- and cross-covariance pattern across regions. Autocovariance patterns are calculated for each sample by embedding it into a time window spanning ± 7 samples and subsequently reducing its dimensionality by selecting a subset of principal components (2 x number of parcels) inferred with a PCA. The dimensionality reduction reduces computational demands and prevents overfitting. To further reduce computational costs, we employed a stochastic variational inference approach (Vidaurre et al., 2018a). Importantly, the distinct auto- and cross-covariance patterns for each HMM state can capture different oscillatory activity, including distinct patterns of multi-region spectral power and phase-locking. Each time a state switches “on”, it can be thought of as a transient bursting event of large-scale, oscillatory network.

#### State-Specific Power and Coherence

State-specific power and coherence were calculated for each participant post-hoc using the source reconstructed data and the a-posteriori probability time course of the states inferred by the TDE-HMM. Following (Vidaurre et al., 2016) a multitaper applying 7 Slepian tapers with a window length of 2 seconds was used to extract time-resolved power and coherence estimates between 1 and 45-Hz with a frequency resolution of 0.5 Hz. Time course data were weighted by the posterior state probabilities prior to applying the multitaper, resulting in state-specific power and cross-spectral densities. To visualise large-scale network connectivity patterns, the mean coherence across states was subtracted from the state-specific coherence values. The resulting values were subsequently thresholded at the 97^th^ percentile to highlight the most prominent connections in the respective figures.

#### State Metrics

Temporal metrics were computed to compare large-scale network dynamics between the two groups. These metrics were calculated for each participant using the padded a-posterior probability time courses of the states. The following state dynamics were extracted: fractional occupancy (fraction of total time a state was present), lifetimes (duration of visits to a specific state), interval times (periods between visits to the same state), state rates (number of state visits per second), and motor cortical beta-power change during state visits. State-specific motor cortical beta-power change was obtained by first subtracting the mean power across all states from state-specific power spectra and then averaging the 18 to 25-Hz power in the superior motor parcels.

#### Robustness Check

The number of states extracted by the HMM is a parameter that must be specified before running the HMM and can yield varying results. Since the inference of the algorithm is randomly initialised, different runs of an HMM with same parameter settings on the same data can yield slightly different results. However, the main state features such as state topologies and dynamics, as well as key findings of the analysis, should be robust across different HMM runs with same parameter settings and varying numbers of states. To ensure that the results obtained were robust, we ran 10 HMMs inferring 8, 10, and 12 states respectively (see SI Fig. 3). The robustness of key findings was checked across all 30 HMMs (See Fig. 4., SI Fig. 6-8, and SI Fig. 11-12). The representative results shown in the main figures come from an HMM with the lowest variational free energy (an approximation to the Bayesian model evidence) of all HMMs that looked to infer 8 states.

### Motor Beta Bursts in Large-scale Network Context

#### TDE-HMM State and Burst Overlap

To investigate the temporal overlap between conventionally estimated motor beta bursts and the occurrence of TDE-HMM network states (Fig. 5), we calculated the percentage of samples where HMM-state occurrences overlapped with motor beta-burst occurrences. This was achieved by dividing the sum of overlapping samples by the total number of burst samples, allowing to interpret the overlap as the percentage of the overall motor cortex beta-burst occurrences. Significance of the overlap between TDE-HMM States and motor beta-burst occurrences was calculated with non-parametric permutation tests. Empirical TDE-HMM state time courses were randomly shifted for each participant to generate null TDE-HMM state time courses that only differed in their temporal relationship to the empirical state time courses. Overlaps between the motor beta burst on-vs-off time courses and the shifted TDE-HMM state time courses were calculated as described above. Overlaps of motor beta bursts with the shifted TDE-HMM state time courses were contrasted against overlaps of bursts with the inferred TDE-HMM state time course with a directed t-test per TDE-HMM state. Bonferroni correction was applied to correct for multiple comparisons across states.

#### Calculation of Network-Associated Beta-burst (NABB) Metrics

We segmented conventional motor beta bursts according to co-occurring large-scale networks derived in the TDE-HMM analysis, to investigate whether focusing on motor beta bursts associated with sensorimotor network activations recovered significant group differences in burst dynamics (Fig. 5A). We refer to these segmented motor beta bursts as Network-Associated Beta Bursts (NABBs). Metrics describing NABB dynamics were calculated in a similar way as the TDE-HMM state metrics. To test whether group differences between individuals with PD and HCs only occurred for sensorimotor NABBs, we identified bursts overlapping with large-scale networks other than the sensorimotor network (other NABBs) and compared their fractional occupancies between the two groups. We further calculated NABB metrics for each large-scale network and compared them between the two groups.

### Statistical Analyses

Group contrasts for Burst, Network-State, and NABB metrics were assessed using GLMs with group constants, age, sex, education, and handedness as regressors to control for potential confounds (SI Fig. 1). Significance was assessed using maximum t-statistic permutation tests by shuffling group labels before calculating the above-mentioned GLMs for 10,000 times. For group contrasts of large-scale network and NABB metrics, where a separate contrast is performed for each network, maximum t-statistic pooling across networks was used to determine significance while controlling for multiple comparisons (Nichols and Holmes, 2002): For each permutation the largest t-statistic across all states was selected to construct a single null-distribution which was used to determine the significance of the metric’s group contrast for each states. Observed t-statistics of state or NABB metrics were deemed to be significant if they exceeded the p < .05 threshold obtained from the null distribution.

UPDRS motor scale items were combined into Bradykinesia/Rigidity and Tremor scores (Hammond et al., 2007; Litvak et al., 2011). Relationships between metrics and UPDRS scores were assessed with GLMs, for all metrics that significantly differed between the two groups. In these models metric scores were predicted from Bradykinesia/Rigidity and Tremor scores while controlling for the above mentioned confounds (SI Fig. 2). Maximum t-statistic permutation tests with 10,000 permutations were applied to determine the significance of the associations.

### Data availability

The conditions of our ethics approval do not permit public archiving of the data supporting this study. Sharing data requires a formal data-sharing agreement in accordance with ethics procedures governing the re-use of sensitive data. Readers seeking access to the data should contact the first author.

## Results

### Decreased static beta power in PD but no differences in conventional beta-burst analysis

The group comparison of motor cortical mean static power spectra (i.e., averaged over the whole resting-state scan) between the HC and PD group (Fig. 2A) yielded a significant cluster in the beta range (18 to 25-Hz) with decreased power in the PD group (*p* < .05). Another non-significant 6 to 9-Hz cluster with increased power in individuals with PD was found as well (*p* = .09). Beta power averaged across the range of the significant cluster was not associated with Bradykinesia/Rigidity (*t*(14) = 0.35, *p* = .72) or Tremor (*t*(14) = 0.06, *p* = .95) scores.

**Figure 2.**
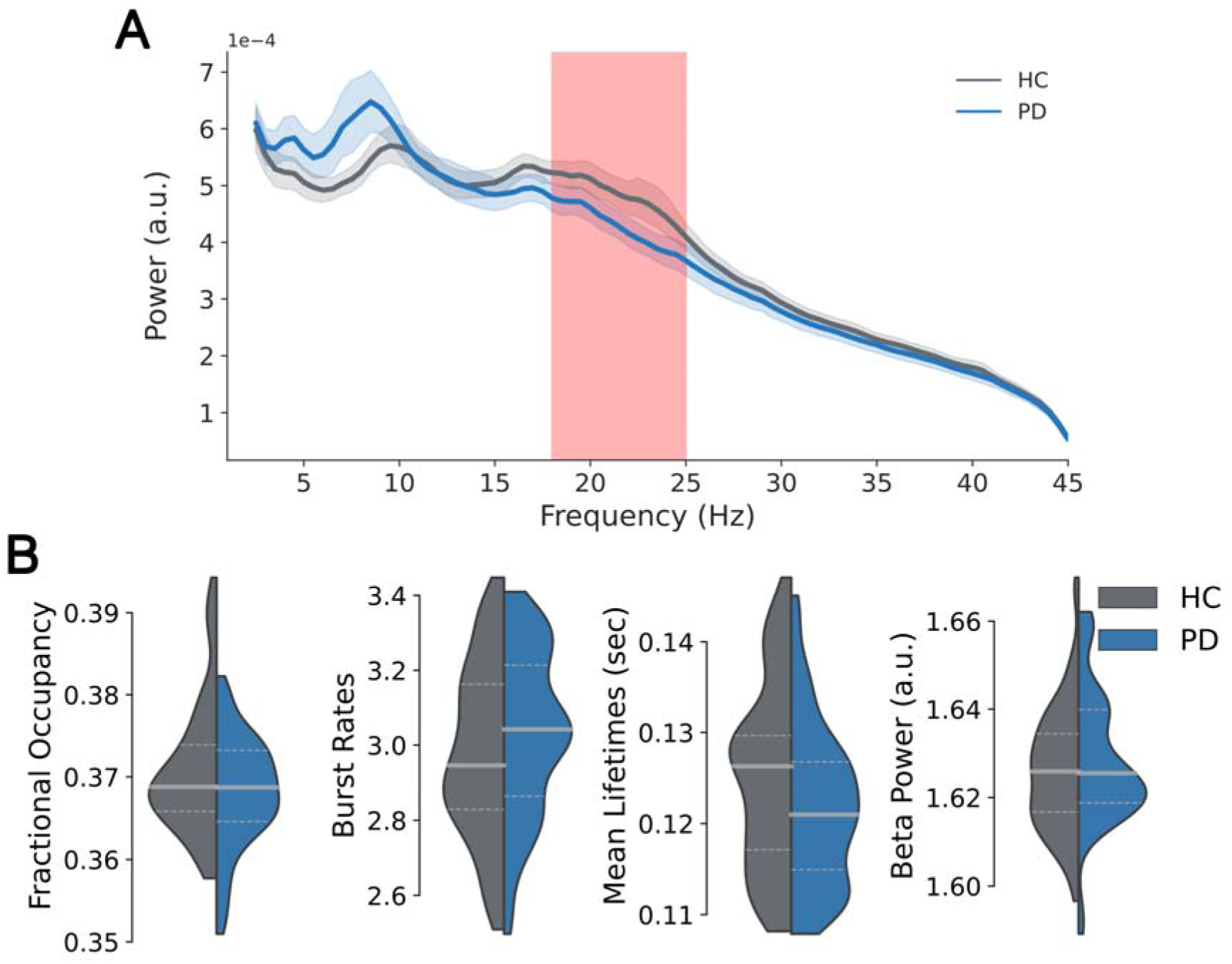
Motor Cortical Beta Oscillatory power decreased in individuals with PD in the static power spectrum, but there were no changes in conventional amplitude-thresholding beta-burst analysis. **(A)** Motor Cortical average power spectra of HC and PD group. Cluster with significant power differences marked in red. **(B)** 18 to 25-Hz motor cortex beta-burst metric group comparison between the HC and PD group. ** indicates p < .01 and * p < .05.

Comparisons of conventional motor cortical beta-burst metrics between the HC and PD group (Fig. 2B) did not reveal significant changes in metrics describing the dynamic properties of bursts (all *p* > .16).

### Hidden Markov Models describe fast, transient network dynamics

Descriptions of temporally and spectrally resolved large-scale networks inferred by the TDE-HMM are depicted in Figure 3. Each time a state switches “on”, it can be thought of as a transient bursting event of distinct oscillatory network activity. These networks exhibited power covariance and phase connectivity patterns consistent with previous studies extracting large-scale networks from MEG resting-state data (Higgins et al., 2021; Vidaurre et al., 2018b). Importantly, State 1 occurrences were linked to an increase in the wideband power and coherence in motor cortical areas. Therefore, from here on, we will refer to State 1 as the sensorimotor state. Since changes in motor cortical dynamics have been reported in PD (O’Keeffe et al., 2020; Pauls et al., 2022; Vinding et al., 2020), we will focus on the sensorimotor state in the following analyses.

**Figure 3.**
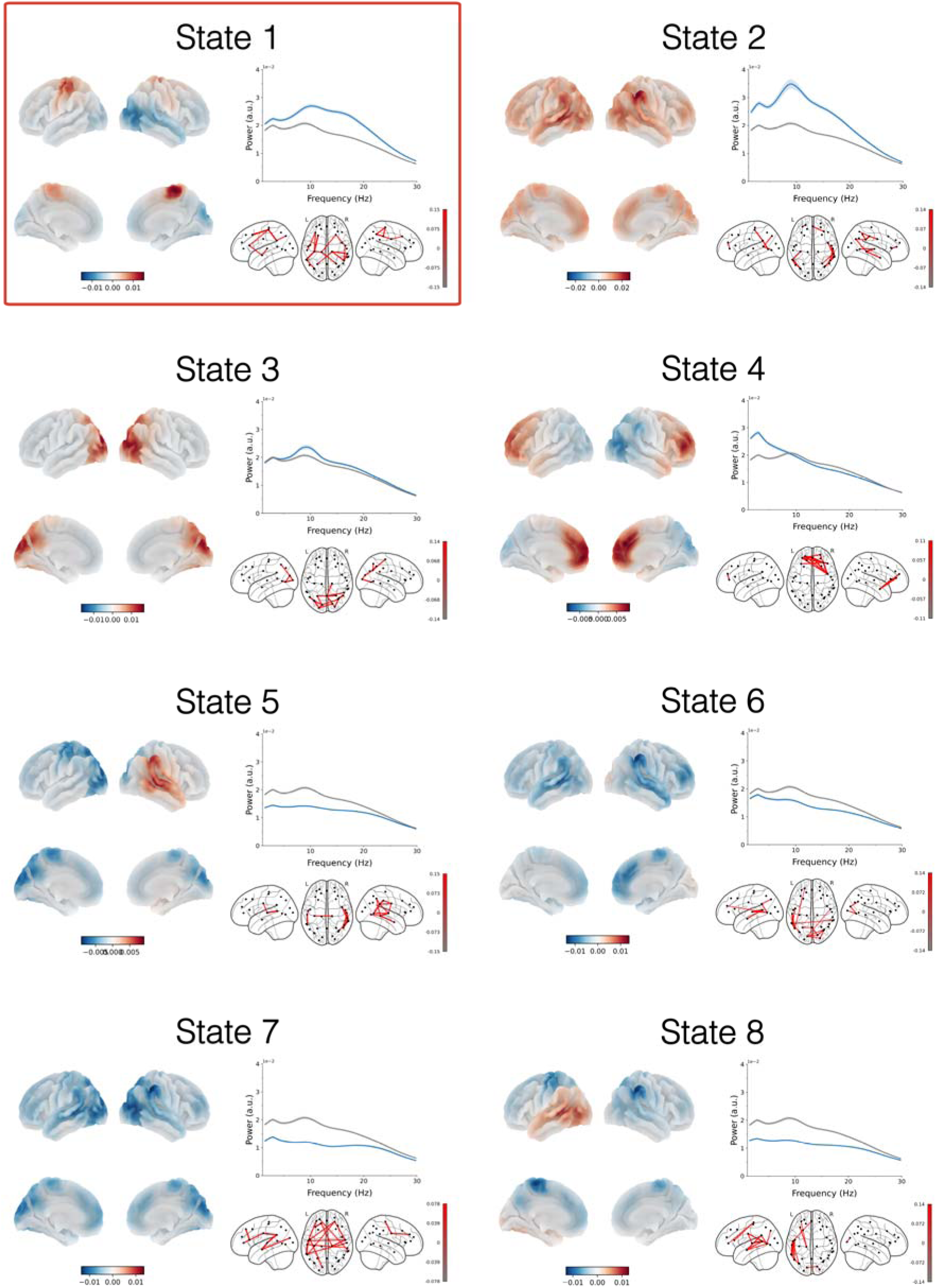
Description of the dynamically switching large-scale networks inferred with TDE-HMM. The red box highlights State 1 as the sensorimotor network state. States are sorted in descending order according to their 18 to 25-Hz motor power. State-specific spatial maps of oscillatory power averaged over 2-30 Hz (relative to the average power across all states) are shown on cortical surfaces. State-specific motor cortical power spectra (blue line) are shown alongside power spectra averaged across all states (grey line). State-specific coherence networks averaged over 2-20Hz (relative to the mean coherence across all states) are shown below the state-specific motor power spectra.

The TDE-HMM state metrics (SI Fig. 4) provide a summary of the temporal dynamics of the large-scale networks captured by the TDE-HMM. Extracted networks had similar mean fractional occupancies, ranging between 11 to 16%. Fractional occupancy values across all states and participants were smaller than 47% and larger than 3% indicating that the brain networks mix well within participants, i.e., all participants’ data were not represented by a single state, and all networks were visited for each participant. Mean lifetimes varied between 50 and 90 ms and interval times between 460 and 778 ms, which is in line with previous studies (Higgins et al., 2021; Vidaurre et al., 2018b).

### Fast, transient network dynamics relate to individual differences in static spectral power

We next assessed whether brain-network probabilities related to static spectral power by correlating fractional occupancies (FOs) of the sensorimotor network with motor cortical beta power across participants (Fig. 4A). As expected, increased occurrence (i.e., FO) of the sensorimotor network was associated with increased beta power, indicating its contribution to the observed power variations across participants. To estimate how network occurrences influence power in a wide range of frequencies, GLMs predicting individual participants’ power spectra from the sensorimotor network probabilities were fitted, and predictions of the obtained models for varying network fractional occupancies extracted (Fig. 4B). Note that a different GLM was fitted for each frequency. Predictions for fractional occupancies of the sensorimotor state indicated a pronounced increase in alpha and beta power in motor cortical areas.

**Figure 4.**
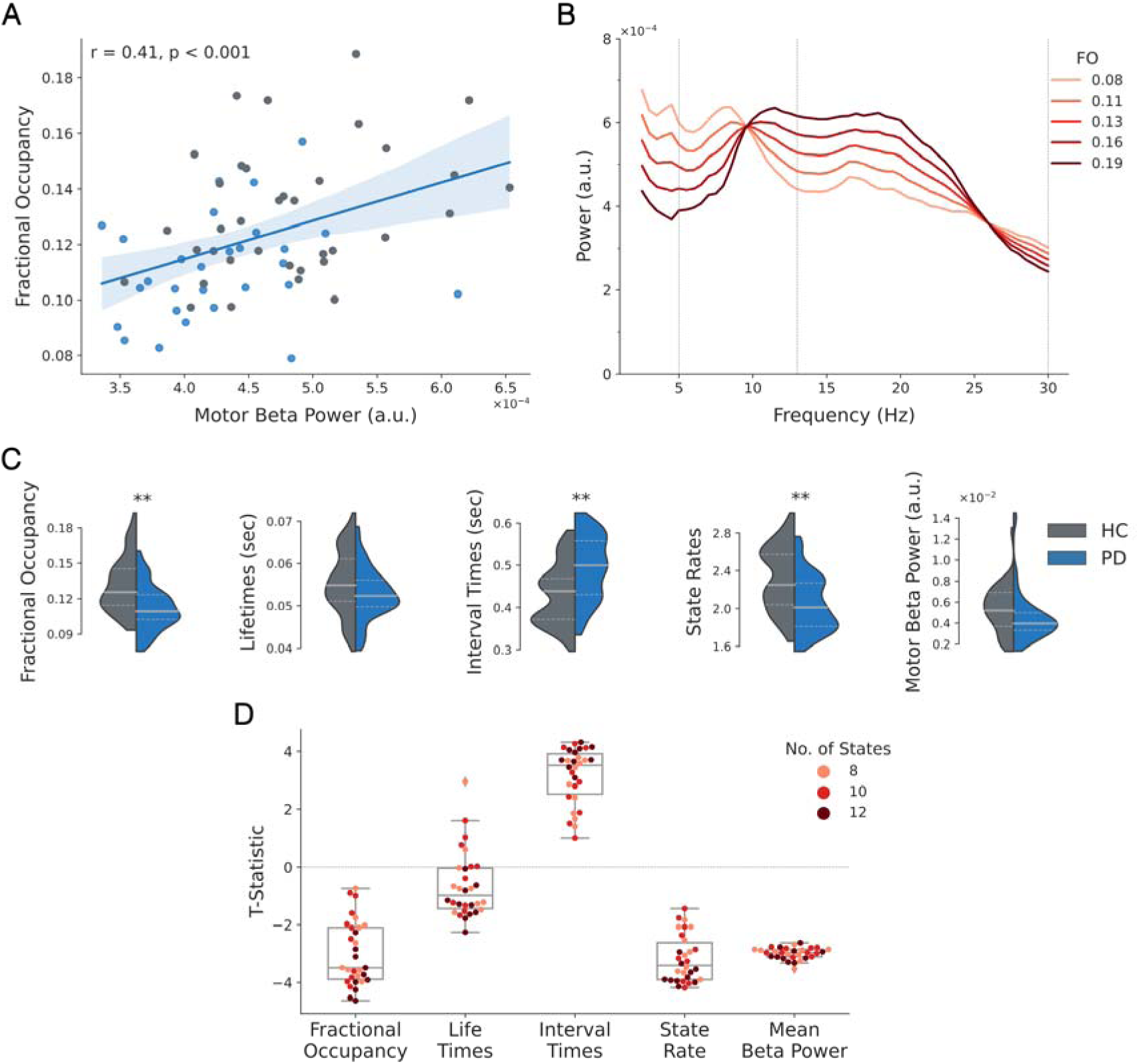
Sensorimotor network dynamics are associated with motor cortical beta power and differ significantly between HC and PD group. **(A)** The significant correlation between each participant’s fractional occupancy of the sensorimotor state and motor cortical beta power confirms that brain state metrics related back to static oscillatory activity. Each dot represents the values of one participant and the colour indicates the participant’s group with HCs coloured in grey and individuals with PD in blue. **(B)** Projections of a GLM that predicts how spectral power in motor cortex varies over subjects, using the fractional occupancy (FO) of the sensorimotor state for each subject. Note that a different GLM was fitted for each frequency. This indicates that as the rate of occurrence (i.e., the FO) of the sensorimotor network increases, so does the alpha and beta power in motor areas. (C) Fractional occupancies, mean lifetimes, mean interval times, state rates, and motor beta power of the sensorimotor network are presented for HCs (grey) and individuals with PD (blue). Significance of group differences is assessed with maximum t-statistic permutation tests controlling for multiple comparisons across states. T-statistics were calculated from GLMs accounting for confounds. (D) T-statistics of significant group differences in state metrics inferred with the TDE-HMM model with 8 States and lowest free energy are robust across 30 model fits extracting the 8, 10, or 12 states. Each dot represents the t-statistic of a group contrast from one of the fitted TDE-HMMs. ** indicates p < .01 and * p < .05.

### Dynamics of sensorimotor network are associated with diagnostic group but not symptom severity

Group comparisons of large-scale network dynamics revealed that the sensorimotor network occurred less frequently in individuals with PD. Contrasts between state metrics of the HC and PD groups (Fig. 4C-G) indicated that the fractional occupancies (*t*(58) = −3.77, *p* < .01) and state rates (*t*(58) = −3.47, *p* < .01) of the sensorimotor state were significantly decreased, whereas interval times were significantly increased (*t*(58) = 3.58, *p* < .01), in the PD group. Additionally, increased lifetimes of a right temporal network (State 8) were observed in PD (*t*(58) = 2.8, *p* = .04). Group comparisons of all other state metric-x-brain state combinations did not reach significance (all *p* > .13) (see SI Fig. 5). In all these analyses, we corrected for multiple comparisons across states for each metric separately with maximum T-statistic permutation tests. Repeating this analysis for 29 other HMMs extracting 8, 10, or 12 States, demonstrated that the observed group differences in the sensorimotor network were robust across different HMM inferences and number of states (Fig. 4H). In the following analyses, we focused solely on the sensorimotor network, because group differences in the right temporal network were not robust across different HMM inferences and parameter selections (SI Fig. 6).

Correlations between sensorimotor state metrics, which differed significantly between the HC and PD groups, and motor symptom severity scores were assessed with GLMs accounting for potential confounds. No significant associations between sensorimotor state metrics of PD participants and Bradykinesia/Rigidity (all *p* > .37) or Tremor scores were observed (all *p* > .17) (SI Fig. 7).

### Motor cortex beta bursts are associated with multiple large-scale networks

We next compared the TDE-HMM network state time courses and conventional motor beta burst on-vs-off time courses (Fig. 5). This revealed that the occurrence of beta bursts overlapped with visits to the sensorimotor state (*t*(132) = 7.47; *p* < .001). This indicated that both analyses were sensitive to similar events in the motor cortex. However, motor beta-burst occurrences also significantly overlapped with State 2 (*t*(132) = 5.44, *p* < .001), which is a state linked to widespread, increased broadband power (Fig. 5B). This suggested that, while motor cortex beta bursts can be associated with the sensorimotor networks, they can also be associated with other networks.

**Figure 5.**
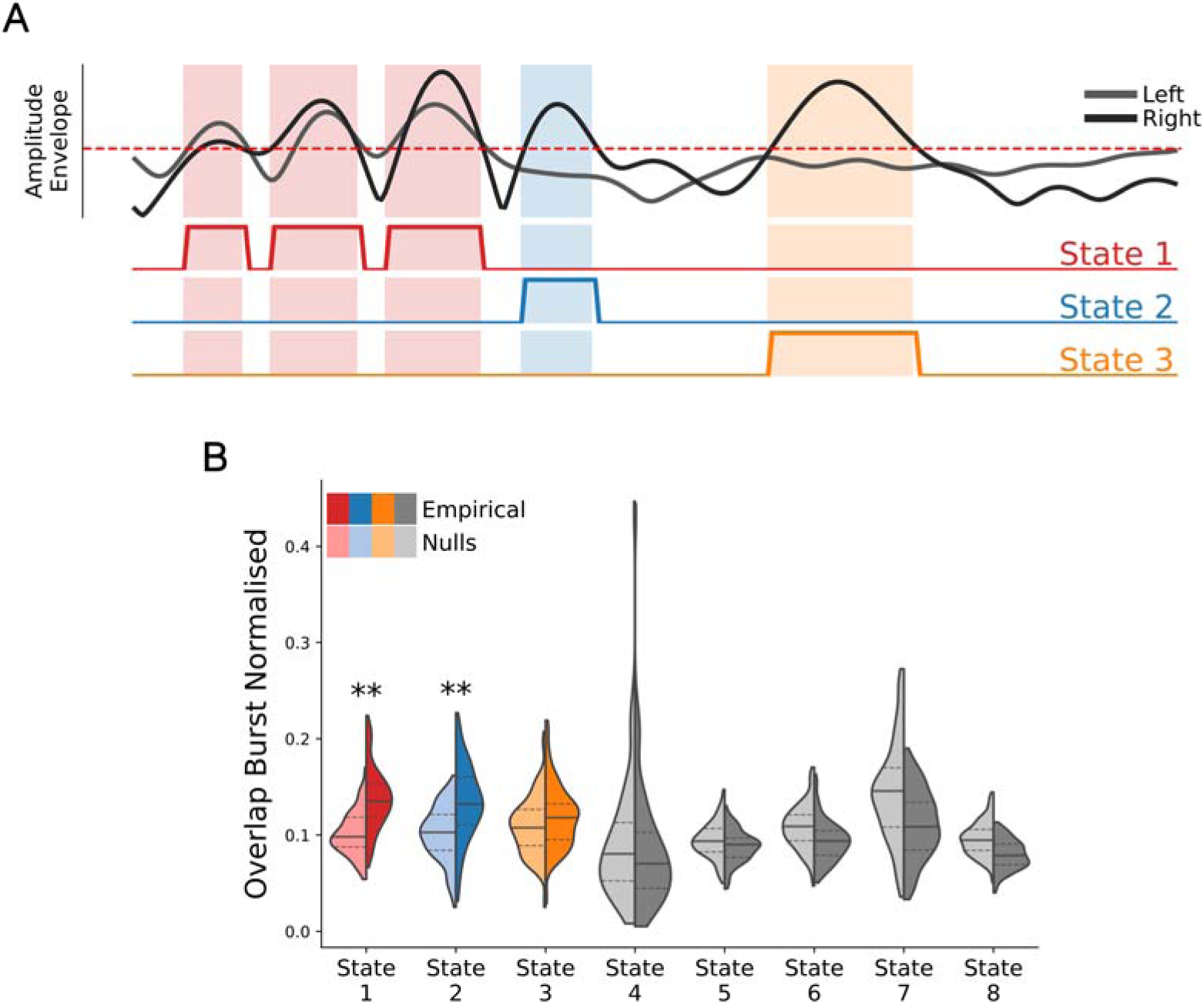
Motor cortex beta bursts extracted with amplitude-envelope thresholding approach do not solely co-occur with sensorimotor network activations. **(A)** Schematic depicting how overlap between the occurrence of conventional beta bursts and HMM network state visits is calculated. Amplitude envelopes of the left (grey) and right (black) motor cortex are thresholded using the 75^th^-percentile (dashed red line). HMM-State on-vs-off time courses are depicted as red, blue, and yellow lines below. Shaded areas indicate periods in which amplitude envelopes cross the 75^th^-percentile and HMM-state occurrences overlap. **(B)** Empirically observed temporal overlap between the occurrence of conventional beta bursts and the different HMM network states. Significance of the overlap was assessed by contrasting the amount of empirically measured overlap (strong colours) with the amount of overlap obtained in the null case (where there is random amounts of overlap), obtained by shifting the HMM-state on-vs-off time courses by a random number of steps (shaded colours) with directed t-tests. Overlaps of beta bursts with HMM-states that were associated with decreases in motor cortical beta power are depicted in grey colours. Bonferroni correction was applied to account for multiple comparisons across states. ** indicates p < .05 and * p < .01 after Bonferroni correction.

### Sensorimotor Network Associated Beta-burst (NABB) dynamics are sensitive to PD

We found that conventional beta bursts in the motor cortex were not predictive of PD. At the same time, we observed that beta bursts can be associated (through overlapping co-occurrence) with multiple networks, and not just the sensorimotor network. To investigate whether focusing on motor beta bursts associated with sensorimotor network activations reveals significant group differences in burst dynamics, we segmented conventional motor beta bursts according to co-occurring HMM networks into NABBs. For example, in the schematic in Figure 5A the overlaps indicate that the first three beta bursts are State-1 NABBs, the next burst is a State-2 NABB, and the final burst is a State-3 NABB.

We found group differences (PD vs HCs) in fractional occupancy (*t*(58) = −4.24, *p* = .001), interval times (*t*(58) = 4.77, *p* < .001) and state rates (*t*(58) = −4.70, *p* < .001) for sensorimotor NABBs (Fig. 6A). This is similar to what we found with the pure TDE-HMM analysis in Figure 4. In short, by titrating the conventional motor cortex beta bursts according to their co-occurrence with large-scale networks, we recovered the group differences observed in the pure TDE-HMM analysis.

**Figure 6.**
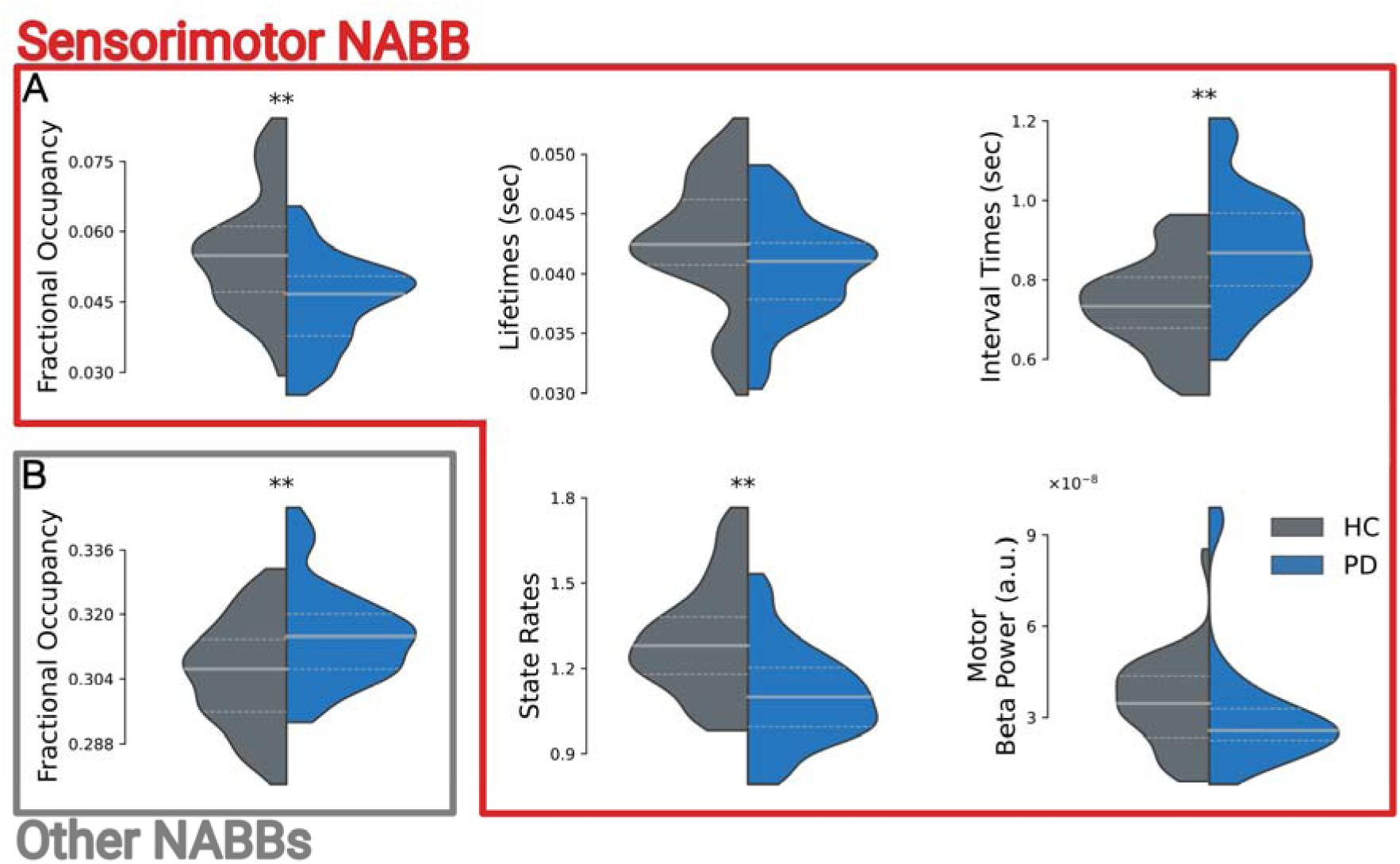
State Metrics of Sensorimotor and frontal network associated beta-burst dynamics (NABBs) significantly differ between HC and PD group. **(A)** Sensorimotor NABB fractional occupancies, mean lifetimes, mean interval times, state rates, and motor beta-power values of HCs (grey) and individuals with PD (blue) are depicted. **(B)** Fractional occupancies of occurrences of all but the sensorimotor NABBs collapsed per group. Significance of group differences is assessed with maximum t-statistic permutation tests controlling for multiple comparisons across states. T-statistics were calculated from GLMs accounting for confounds. ** indicate p < .01 and * p < .05.

To further test whether PD-related decreases in sensorimotor NABBs were specific for the sensorimotor network, we calculated the probability of bursts to overlap with any of the other networks (other NABBs) for the two groups. We observed that these other NABBs were significantly more likely to happen in the PD group (*t*(58) = 3.18, *p* = .005; Fig. 6B). This suggests that the decrease in sensorimotor NABBs may be attenuated by activity in other NABBs in conventional burst analyses that ignore the network context of motor beta bursts. In follow-up analyses investigating whether certain NABBs drive the PD-associated significant increase in the other NABBs analysis, we found only State-7 NABBs to be significantly more likely (*t*(58) = 3.01, *p* = .02) and to show longer lifetimes in individuals with PD (*t*(58) = −2.98, *p* = .03) (see SI Fig. 9-12). All other NABBs did not significantly differ between the two groups.

## Discussion

In the present study, we investigated whether analysing motor cortical activations within their large-scale brain network contexts provides a more sensitive measure of motor cortical, oscillatory changes in individuals with PD. The static power-spectrum approach showed decreases in motor cortical beta power in the PD group. Conventionally estimated motor beta-burst dynamics did not differ between the two groups, whereas large-scale network dynamics obtained with a TDE-HMM revealed decreases in the probability of the sensorimotor large-scale network being present. We hypothesised that the differing findings may be due to different levels of network specificity of these two approaches. Accordingly, we demonstrated that conventional motor cortex beta bursts did not solely co-occur with activation of the sensorimotor network, but also with more widespread network activations. Separating motor cortex beta bursts co-occurring with sensorimotor network activations from bursts co-occurring with activations of other large-scale networks allowed us to recover significant group differences between the HC and PD group in burst visits, similar to what was observed in the pure large-scale network analysis. Importantly, group differences between beta bursts co-occurring with sensorimotor network visits and beta bursts co-occurring with other large-scale networks were in opposing directions, suggesting that these opposing changes may diminish group contrasts in conventional burst analyses that ignore the network context of motor beta bursts. Taken together, we replicated classical motor network results, provided context about why they might be inconsistent in the non-invasive literature, and highlighted the importance of taking a whole-brain focused perspectives to precisely describe changes in cortical oscillatory activity in PD.

### Static motor cortex focused analysis reveals changes in oscillatory activity

The static power-spectrum analysis revealed significant decreases in motor cortical beta power. In line with this, decreased motor cortical beta power in PD (Heinrichs-Graham et al., 2014; Vardy et al., 2011) and increased beta power after administration of dopaminergic medication (Cao et al., 2020; Heinrichs-Graham et al., 2014; Melgari, 2014) have been previously reported. However, it is important to stress that electrophysiological recordings of motor cortical beta oscillations have yielded varying findings (Boon et al., 2019; Underwood and Parr-Brownlie, 2021). Additionally, widespread beta power decreases, potentially linked to spectral slowing, have previously been observed in central and posterior regions of the brain (Bosboom et al., 2006; Stoffers et al., 2007) that likely contribute to the observed beta power decreases. It remains an open question what causes these varying findings on motor cortical beta changes in MEG recordings of individuals with PD. Methodological issues (O’Keeffe et al., 2020), or systematic differences in PD-sample properties across studies, i.e., differing medication status, PD-subtypes, motor status, or disease progressions across studies (Boon et al., 2019), might be potential reasons. Another reason may be that the spatial extent (Zich et al., 2020) or network context of motor cortical high beta-power events needs to be considered to dissect beta events that may be affected by PD in different ways.

### Sensorimotor large-scale network changes in PD group

The dynamics of the sensorimotor network, extracted with the TDE-HMM analysis, indicated a lower probability of this network to be present, reflecting decreases in the occurrences and longer intervals in-between visits. The observed reduction of sensorimotor network occurrences in individuals with PD replicates previous findings using static networks derived from ICA (Schneider et al., 2020), network-based statistics (Yassine et al., 2022) and motor cortical beta bursts (Vinding et al., 2020). Burst rate reductions were reported to be significantly associated with Bradykinesia scores (Vinding et al., 2020), whereas state probability changes observed in our TDE-HMM analysis were not significantly associated with motor symptom severity scores. We linked variations in the fractional occupancies of the sensorimotor network to variations in static beta power, suggesting that these changes in large-scale network dynamics underpin static power-spectrum measures. In sum, our findings indicate that large-scale network descriptions of brain activity enable distinguishing between individuals with PD and HCs and shed light onto the dynamic underpinnings of group differences in the static power spectrum.

Importantly, extrapolating results from studies investigating the role of beta activity in the basal ganglia-thalamocortical loop through invasive recordings of PD patients, one would hypothesise that motor cortical beta power should be increased in PD patients. This is in contradiction to the varying findings on motor cortical beta power changes reported in non-invasive M/EEG studies (Boon et al., 2019) as well as to our specific observations of decreased beta power and sensorimotor network occurrence probability. Several factors could account for this surface-level inconstancy between invasive and M/EEG studies: 1) Volunteers in invasive ECoG-or DBS-studies tend to have more severe disease progressions, symptomatology, exposure to and ‘wearing off’ of dopaminergic medication; and they may be less heterogenous due to surgery qualification criteria. 2) Comparisons of on-vs-off medication, prevalent in invasive studies are not equivalent to comparisons of PD-vs-HC groups, prevalent in non-invasive studies. 3) Due to the larger scale of measurement, M/EEG may pick up changes in oscillatory power due to other factors related to PD pathology, e.g., oscillatory slowing, which dilute basal ganglia-thalamocortical loop related motor cortical activity in MEG recordings. Studies systematically investigating the effect of these factors on the inconsistencies between invasive and non-invasive studies may help to improve our understanding on how findings of these modalities related to each other. This in turn would enable drawing better conclusions about PD-related pathologies that might underpin non-invasive electrophysiological biomarkers.

### Network context identifies PD-related changes in cortical beta-burst

In contrast to conventional amplitude-thresholding burst analyses, the spatiotemporal nature of large-scale networks allows for increased confidence about the spatial extent of network activation at each moment. We show that it is crucial to consider the underlying network distributions of transient, high amplitude beta events when trying to identify sensorimotor network specific markers of PD or other disorders. In contrast to previous reports (O’Keeffe et al., 2020; Pauls et al., 2022; Vinding et al., 2021, 2020), we did not initially observe changes in motor burst metrics in individuals with PD. Instead, we demonstrated that our conventional amplitude-threshold beta-burst analysis did not only identify bursts co-occurring with sensorimotor network activity, but also during occurrences of more widespread network activations. By separating motor beta bursts co-occurring with the sensorimotor network from motor beta bursts co-active with other large-scale networks, we revealed significantly decreased fractional occupancies and bursts rates in motor beta bursts of individuals with PD, similar to what has been reported by Vinding et al. (2020). These findings suggest that a large-scale network perspective enables for a more precise description of PD-related changes in sensorimotor network activity.

Interestingly, the group difference t-statistics obtained from the sensorimotor NABB metrics were larger than the t-statistics obtained from the sensorimotor network state metrics from the pure HMM analysis. This might indicate that targeting sensorimotor network visits with high motor cortical amplitudes can further refine predictability of PD-related changes in motor cortical activity.

Motor beta bursts associated with different large-scale networks, showed differing patterns of change in the rate of beta-burst occurrence. In contrast to sensorimotor NABBs, co-occurrence probabilities of beta bursts with all other large-scale networks (combined) were significantly increased in individuals with PD. This indicates that analysing dynamics of all motor cortex beta bursts together, without subdividing them according to networks with which they are most associated, attenuates group differences. Thus, it may be crucial to extract sensorimotor networks or sensorimotor NABBs using the large-scale network context of motor beta bursts to reliably differentiate between HCs and individuals with PD.

### Putative contributions of NABBs

Further, investigating which of the non-sensorimotor NABBs is driving the increases in the PD group in the other NABBs analysis revealed that State 7 NABBs played a significant role, being more likely to be present and having significantly longer lifetimes. Strikingly, the increase of lifetimes of the State 7 NABBs agrees with findings of increased burst lifetimes by Pauls et al. (2022), whereas the decrease in sensorimotor NABB rates is in line with reports of decreased burst rates by Vinding et al. (2020) Thus, it is tempting to speculate that by embedding beta bursts in simultaneously occurring large-scale networks, different beta bursts can be disentangled. Differences in the activation of these NABBs in different PD subtypes may have the potential to reconcile differing findings in the literature.

Interestingly, increased State 7 NABB lifetimes match burst dynamics expected based on invasive studies of the basal ganglia-thalamocortical loop (Tinkhauser et al., 2018). Thus, they may provide a dynamic window in which basal ganglia-thalamocortical loop related activity can be picked up in M/EEG recordings of motor cortex. If this hypothesis holds, basal ganglia-thalamocortical projections would not be associated with the sensorimotor network but rather with a state of decreased oscillatory activity. This hypothesis should be treated carefully as this analysis was exploratory and requires replication in other datasets. Studies investigating the dynamics of STN-motor cortical connectivity and its cortical network context could test this hypothesis.

### Limitations

Our sensitivity to link neural changes to motor symptoms may have been compromised by using bilateral measures. All cortical measures that were linked to PD motor-symptom severity were collapsed across both hemispheres. Similarly, we used Bradykinesia/Rigidity scores, calculate from UPDRS-III item scores for both hemi-sides, as an approximation for motor symptom severity of individuals with PD (Hammond et al., 2007; Litvak et al., 2011). In the context of PD, where the expression of motor symptoms is often asymmetric, this might reduce the sensitivity to unilateral associations between neural measures and motor symptom severity scores.

The spatial resolution of the used parcellation is limited. All our analyses were conducted on source localised data based on a weighted parcellation with 39 cortical regions (Colclough et al., 2015). Parcels in this parcellation are derived from a group spatial ICA of fMRI recordings from the human connectome project. Since several motor areas, i.e., Supplementary Motor Area (SMA), pre-SMA, or motor cortex, are represented by the same parcels, no conclusions about different processes in these areas can be made. Refinement of the parcellation may allow for more precise claims about areas involved in the respective networks.

### Summary

In sum, the present study demonstrated that the spatiotemporal context of motor cortical activations is important for identifying PD-related changes in neuronal activity. We fitted a TDE-HMM to our data, providing a rich spatiotemporal description which can be linked back to the fundamental static power spectrum. The sensorimotor network dynamics distinguished between individuals with PD and HCs, whereas a conventional amplitude thresholding bursts analysis failed to do so. The nature of the TDE-HMM approach provides greater confidence in identifying sensorimotor network beta activity because of the full network dynamics model. Finally, we provided evidence that relating conventionally estimated motor beta bursts to large-scale network activity may be crucial for exploring PD-related changes in the sensorimotor network and may help to clarify the contradicting findings in the literature. This work highlights the importance of taking a large-scale network perspective when attempting to identify non-invasive cortical biomarkers for PD.

## CRediT authorship contribution statement

O.K., N.Z., A.C.N., and A.Q. designed the study; N.Z. recorded the data; O.K. curated the data; O.K., C.G, and A.Q. performed formal data analyses; O.K., C.G., A.Q., A.C.N., and M.W. interpreted the data; O.K., C.G., M.T.H., A.C.N., M.W., and A.Q. wrote the paper; and A.C.N., M.W., and A.Q. supervised this project.

## Abbreviations

ECoG: Electrocorticogram
EEG: Electroencephalogram
HC: Healthy Control
ICA: Independent Component Analysis
MEG: Magnetoencephalogram
NABB: Network-associated Beta Bursts
PD: Parkinson’s Disease
PCA: Principal Component Analysis
SD: Standard Deviation
STN: Subthalamic Nucleus
TDE-HMM: Time-delay Embedded Hidden Markov Model
UPDRS: Unified Parkinson’s disease Rating Scale

