## Supplementary Information for "Changes in sensorimotor network dynamics in resting-state recordings in Parkinson’s Disease"

Oliver Kohl^1^, Chetan Gohil^1^, Nahid Zokaei^1,2^, Michele T. Hu^3^, Masud Husain^2,3^, Anna C. Nobre^1,2^, Mark Woolrich^1^, and Andrew Quinn^1,4^

Supplementary Material

### Detailed description of combined PD volunteer samples

**SI Table 1 Detailed descriptions of the two PD volunteer samples combined for this study.**

|  | **Zokai et al. (2021)** | **Heideman et al. (2020)** |
| --- | --- | --- |
| N | 12 | 16 |
| Groups matched in | Age  Education | Age  Education |
| Inclusion Criteria | Currently not participation in ongoing drug trial  Not taking any of the following drugs:  Psychotropic hypertensive  Vasoactive medication  Long-acting dopamine agonists  No history of neurological or psychiatry disorders other than PD.  Tolerating coming off dopaminergic medication. | Being diagnosed with PD within the last 5 years.  Being able to understand written and spoken instructions in English.  Being older than 50 years.  Tolerating coming off dopaminergic medication. |
| Dopaminergic Medication | Off-Medication  Since 7 p.m. of previous day. | Off-Medication  Since 7pm of previous day. |
| UPDRS administration | By trained clinician. | By trained clinician. |
| Ethics-Reference | 12/SC/0650 | 12/SC/0650 |
| Cohort | Participants were recruited from neurological clinics in Oxfordshire (UK). | Dementias and Neurodegeneration Speciality (https://dendron.org.uk) |
| Gender  (female/male) | 8/4 | 7/9 |
| Handedness  (right/left) | 11/1 | 14/2 |
| Age (years) (mean, range) | 68.5 (57-77) | 68.5 (54-79) |
| Education (years) (mean, range) | 16 (10-24) | 13.69 (10-23) |
| Years since Diagnosis (years) (mean, range) | 3 (1-7) | 2.75 (1-4) |
| UPDRS III motor score  (mean, range) | 32.33 (20-55) | 28.86 (11-49) |
| Hoehn and Yahr scale (mean, range) | 1.75 (1-3) | 1.78 (1-3) |
| Levodopa-equivalent daily dose (mg/day) (mean, range) | 283.75 (150-540) | 488.39 (300-900)* |

N = number of participants; UPDRS = Unified Parkinson Disease Rating Scale
* Two volunteers with PD did not take Levodopa (or equivalent) medication.

### Supplementary Figures for GLM-Analyses

#### Summary of GLMs used for group comparisons between HCs and PD patients.

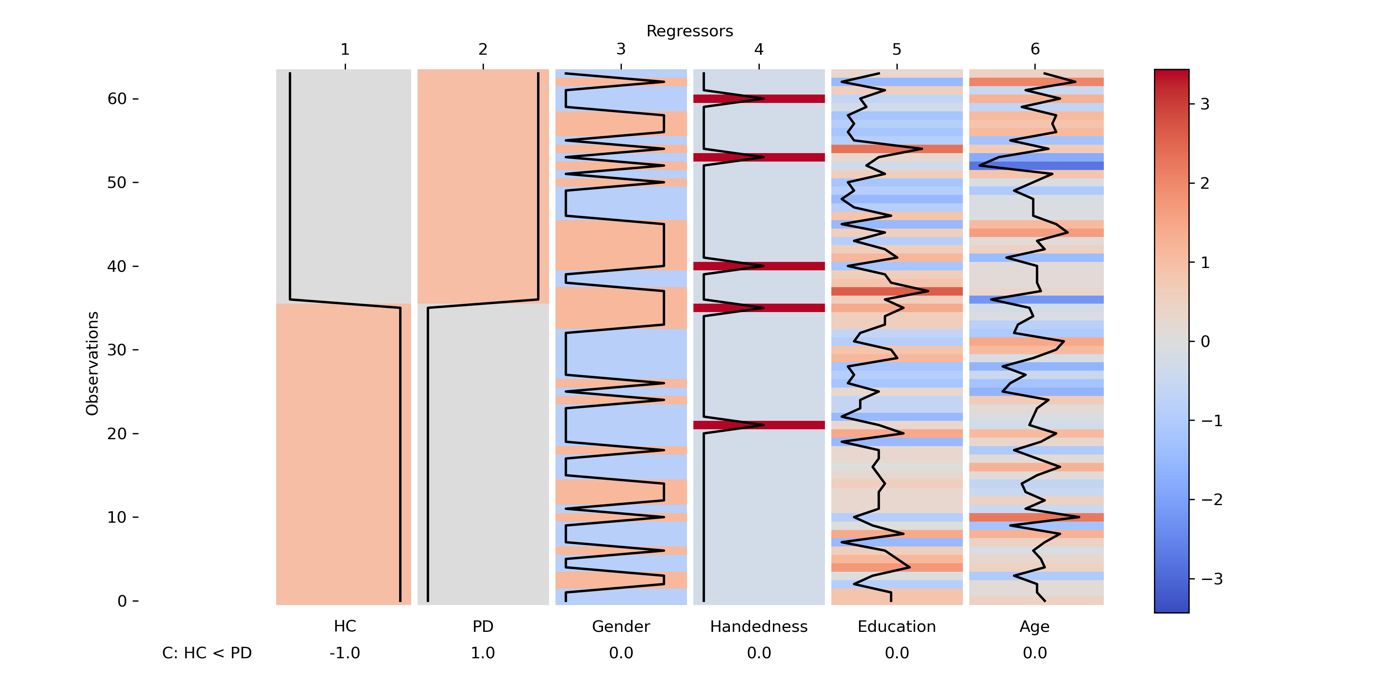

**Figure SI 1 Summary of GLMs used to compare variables between HCs and PD patients.** Regressors added to GLMs are represented as bars with each row representing the z-scored regressor value of a single participant. Contrast between HCs and PD patients is depicted below. Note that the contrast is specified in such a manner that positive t-statistics indicate larger values for PD patients and negative t-statistics larger values for HCs.

#### Summary of GLMs used to associated Variables with Motor Symptom Severity Scores.

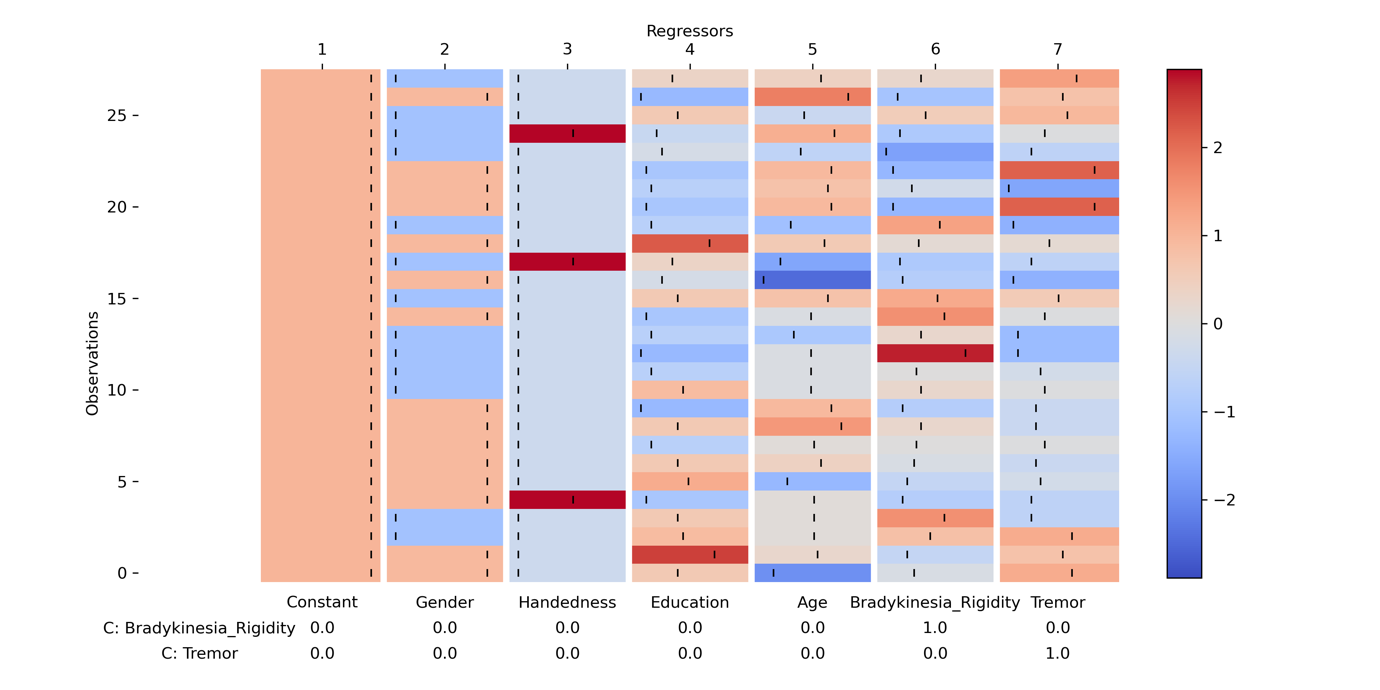

**Figure SI 2 Summary of GLMs used to calculate associations between variables and motor symptom severity scores.** Regressors added to GLMs are represented as bars with each row representing the z-scored regressor value of a single participant.

### Free Energy of different TDE-HMM fits

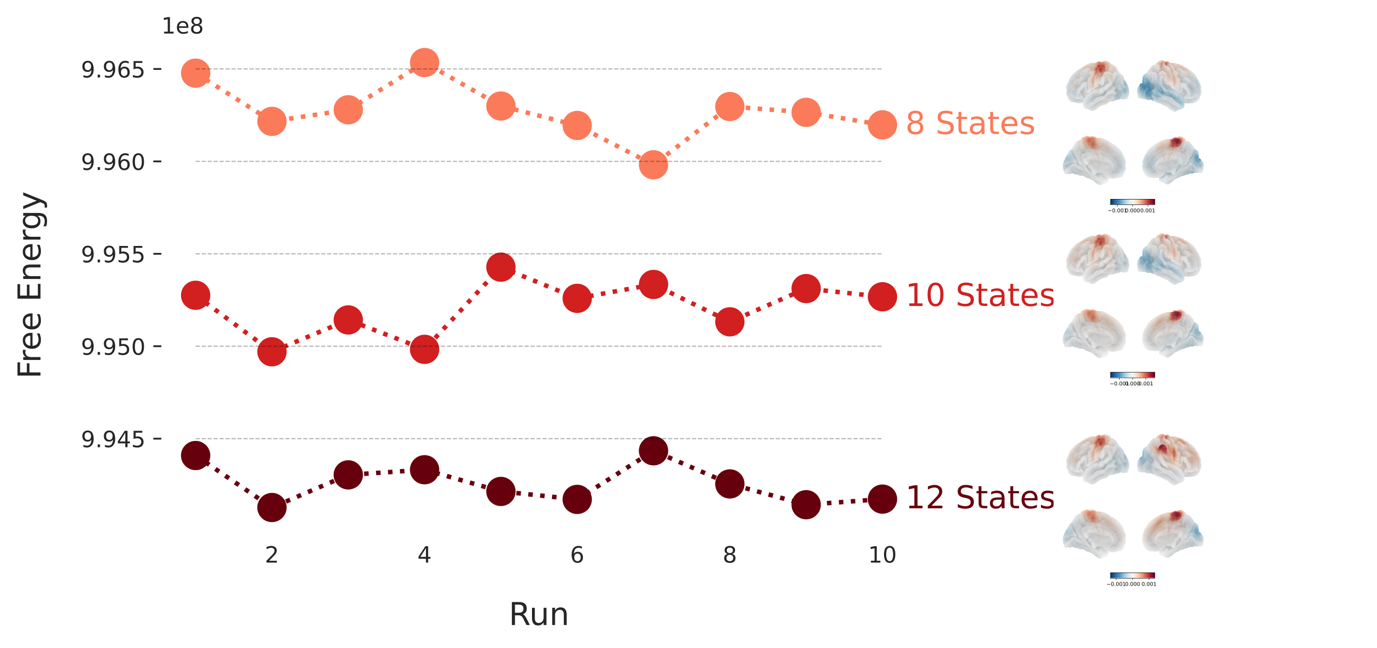

**Figure SI 3 Free Energy values of all HMM fits.** Free energy values for each of the 30 HMM fits are represented by a dot. Colours mark HMMs inferring different numbers of states. Wideband power maps of the sensorimotor network for the HMM fit with the lowest free energy of all HMMs extracting the same number of states are presented on the right.

### HMM-State Metric Analysis

#### State Metrics across all participants

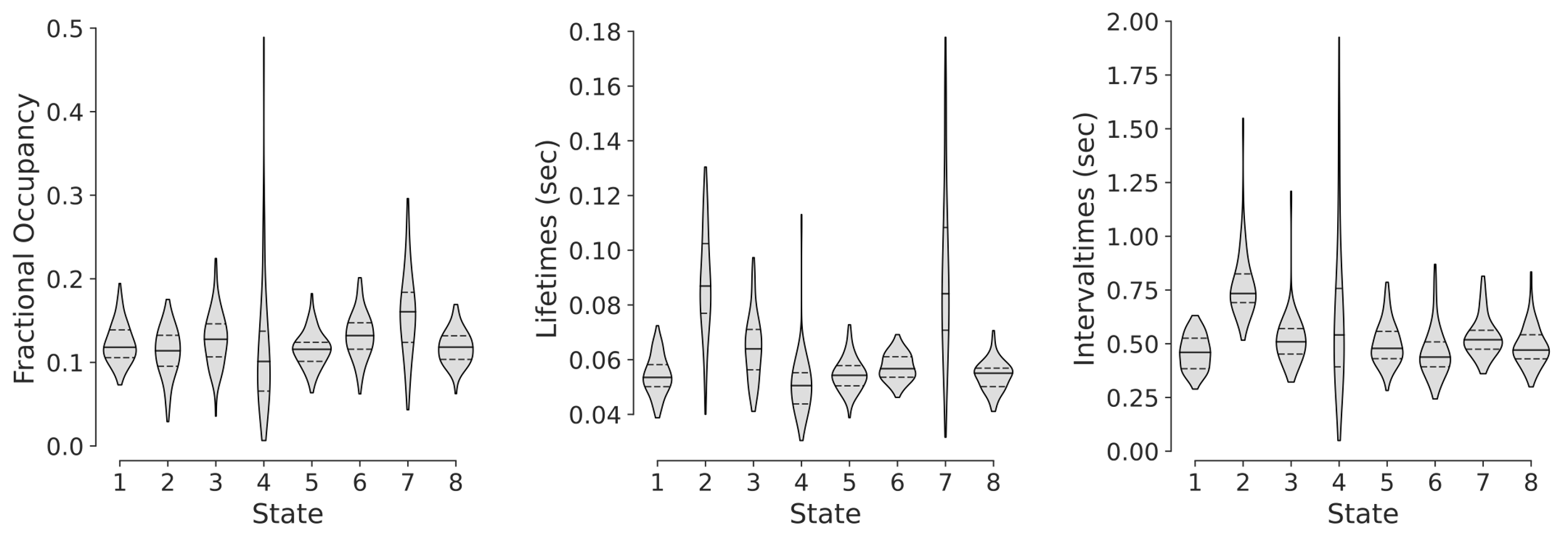

**Figure SI 4 State Metrics of TDE-HMM fit with lowest free energy extracting 8 states.** Fractional Occupancies of all States are below 0.5 and larger than 0 indicating that the extracted HMM-states mix well within and between participants.

#### State Metric Group Contrast

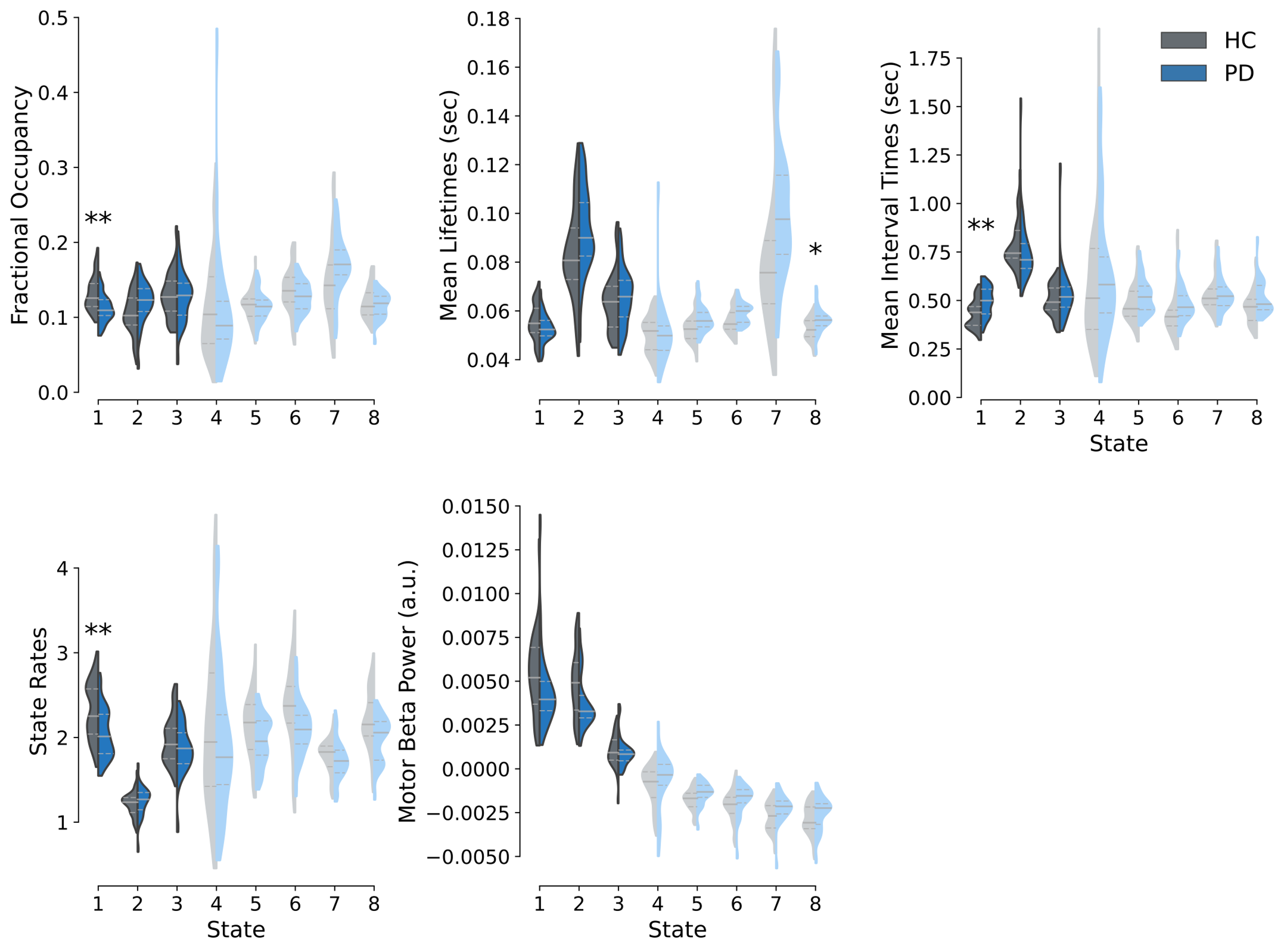

**Figure SI 5 Large-scale network dynamics group contrast between HCs and PD patients.** Fractional occupancies, mean lifetimes, mean interval times, state rates, and motor beta power change of HCs (grey) and PD patients (blue). Significance of group differences is assessed with maximum t-statistic permutation tests controlling for multiple comparisons across states. T-statistics were calculated from GLMs accounting for confounds. State metrics of large-scale networks whose occurrence results in a decrease of beta power are plotted in shaded colours.
** indicates p < .01 and * p < .05.

### HMM-State Metric Analyses Robustness

#### State Matching Procedure

Analyses calculating TDE-HMM state metric group contrasts and state metric x UPDRS score associations were repeated across all the 30 HMMs to check whether key findings in the reported HMM fit generalise across different initialisations and number of states. Since the initialisation of the TDE-HMM is random, the order of inferred states is arbitrary, and states have to be matched before assessing the robustness of the observed effects. To match states across different runs inferring the same number of states, we first identified the HMM with the lowest free energy and matched states of all the remaining HMMs to it. To match states between 2 HMMs, observation models for all states were extracted and correlation distance between all states of the 2 HMMs runs were calculated. This resulted in a number of states x number of states distance matrix. The Hungarian algorithm was applied to these distance matrices to find an optimal state assignment between the states of the 2 HMMs with a minimal overall distance. This procedure was performed separately for all HMMs inferring 8, 10 or 12 states. To also allow for comparisons between runs with different numbers of states, the states of the HMMs with the lowest free energy extracting 10 and 12 states were matched to the HMM with the lowest free energy inferring 8 states. Correlation distances were calculated between all states of the HMMs resulting in an 8x10 or 8x12 distance matrix. These distance matrices were zero padded to a square shape of either 10x10 or 12x12 so that the Hungarian algorithm could be applied to them. States of the HMMs with 10 or 12 states that were allocated to one of the states of the 8-state HMM were carried on for the robustness check, whereas states matched to one of the zero padded columns were discarded. States of the remaining HMMs inferring 10 or 12 states with higher free energy were first matched to the HMM with the lowest free energy inferring the same number of states, before being matched to the HMM with the lowest free energy inferring 8 states.

#### Robustness of Group Contrast

State metric group contrasts of the sensorimotor state and the left temporal state were repeated for each of the 30 HMMs to assess the robustness of the group differences across different HMM fits. The majority of group contrasts of the sensorimotor’s fractional occupancies, interval times and bursts rates yielded t-statistics larger than 2 because of what we concluded that these group differences are robust across HMM fits. For the group contrast of the left temporal state’s lifetimes however, many HMM fits yielded t-statistics lower than 2 because of what we concluded that lifetime differences in the left temporal state are not robust across different HMM fits (see SI 6).

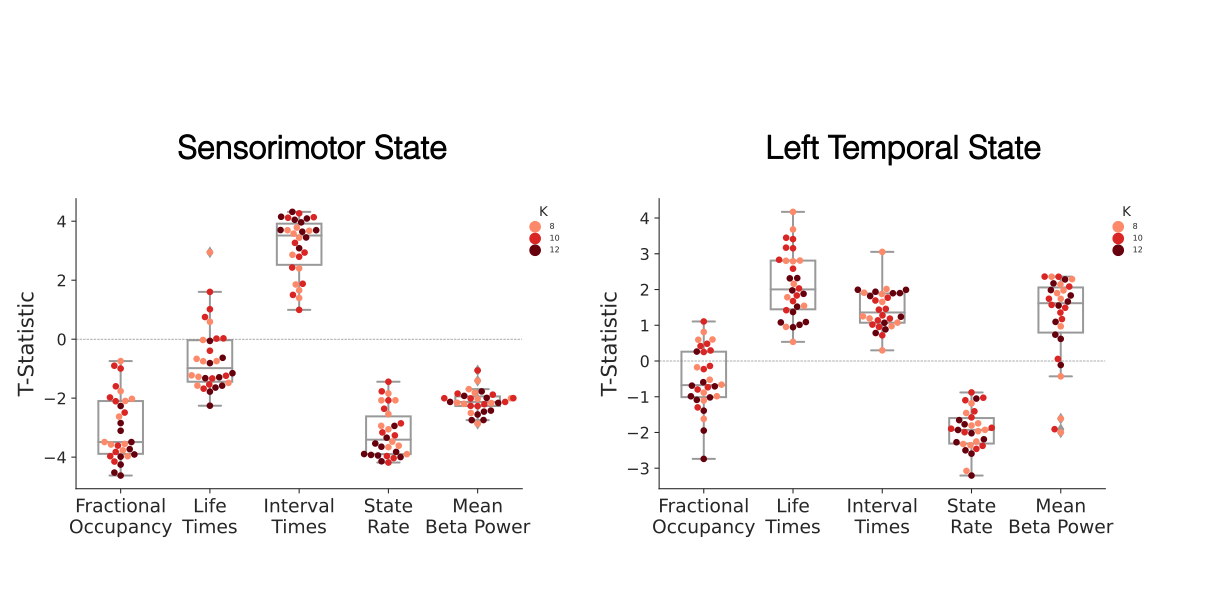

**Figure SI 6 Group differences of sensorimotor network metrics are robust across all HMM inferences**. Each dot represents the t-statistic from the respective state metric’s group contrast between HCs and PD patients for a different HMM inference. The colours of dots indicate how many states were inferred for the depicted HMM inference.

#### Robustness of State Metric x UPDRS score association

Associations between Bradykinesia/Rigidity scores and state metrics of the sensorimotor state were calculated for all 30 HMMs. For both states t-statistics of all associations were smaller than two and closely distributed around 0 indicating that the non-significant associations between Bradykinesia/Rigidity scores and state metrics were robust across all HMM fits (see SI 7).

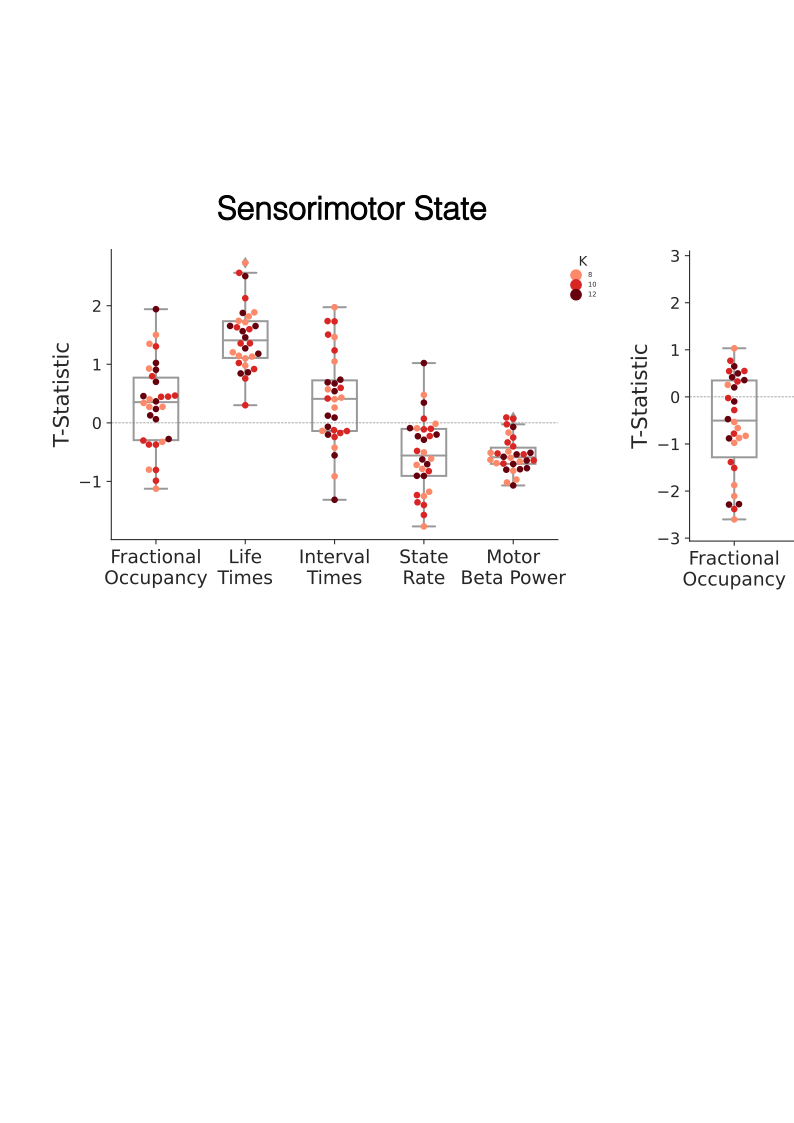

**Figure SI 7 Lack of associations between sensorimotor network metrics and Bradykinesia/Rigidity scores is robust across all HMM inferences.** Each dot represents the t-statistic from the respective state metric’s correlation with Bradykinesia/Rigidity score for a different HMM inference. The colours of dots indicate how many states were inferred for the depicted HMM inference.

### Sensorimotor vs. Other NAABs analysis robustness

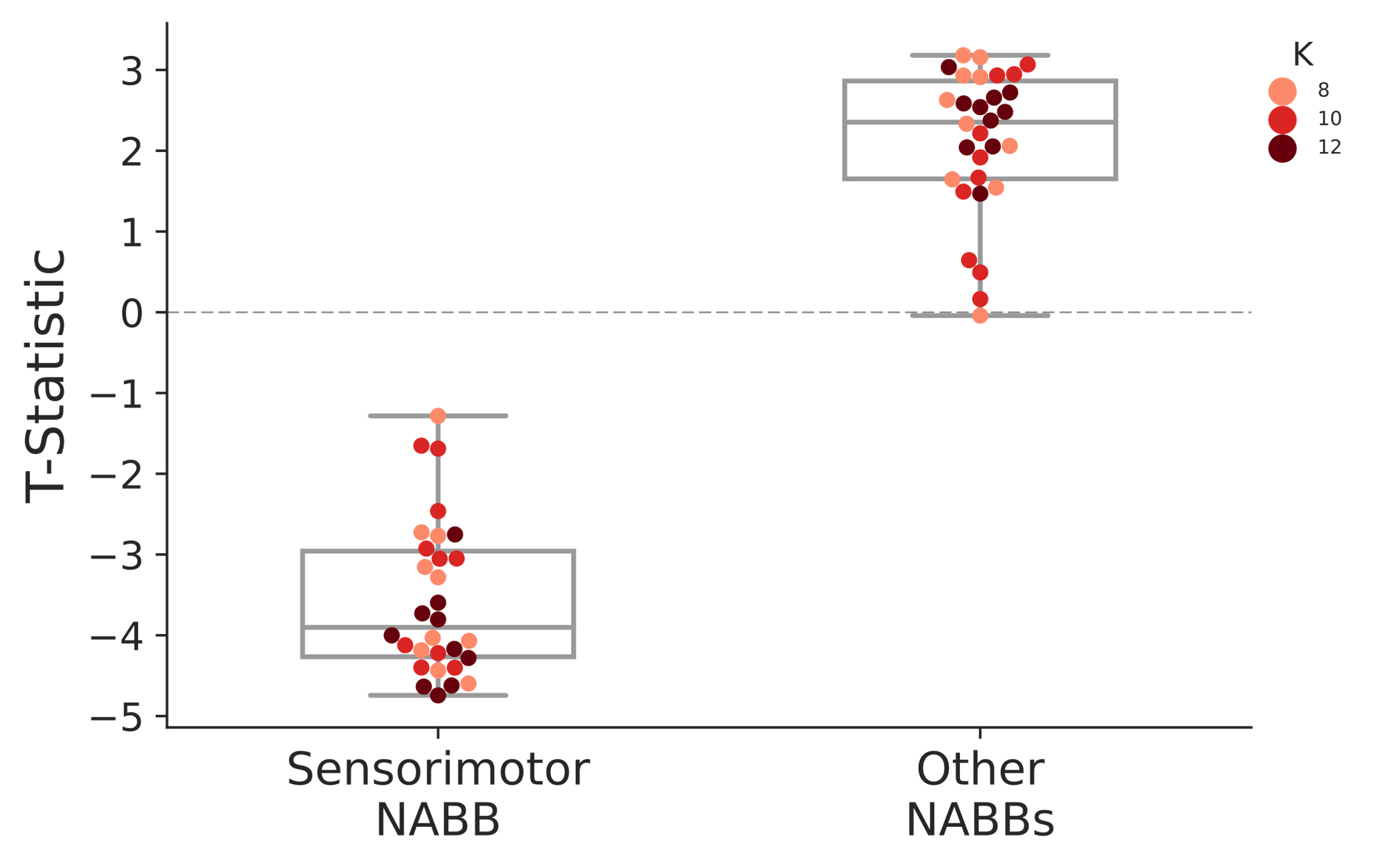

**Figure SI 8 Group differences of sensorimotor and other NABBs fractional occupancies between HCs and PD patients are robust across different HMM-inferences and number of states.** Each dot represents the t-statistic from the respective NABB’s fractional occupancy group contrast for a different HMM inference. The colours of dots indicate how many states were inferred for the depicted HMM inference.

### NABB Metric Analysis

To better understand whether PD-related increases in the other NABBs analyses were due to balanced increases in all but the sensorimotor NABBs or due to increases in specific NABBs, we calculated NABB metrics for each of the obtained large-scale networks.

We observed that co-occurrences between motor cortical beta bursts and State 7 were significantly more likely in PD patients (*t*(58) = 3.01, *p* = .02) and occurred for significantly longer intervals (*t*(58) = -2.98, *p* = .03) (Fig. SI 9). The fact that this increased probability in co-occurrences was correlated with PD patient’s Bradykinesia/Rigidity scores (*t*(58) = 2.2, *p* = .04) and high beta coherence (*r* =.39, *p* = .002) suggests that informing the beta burst analysis with TDE-HMM large-scale network dynamics might reveal beta bursts with different functionalities (SI 10). Repeating the NABB analyses for all 30 TDE-HMM fits (10 fits for 8, 10, and 12 states respectively) demonstrated that the observed group differences in the sensorimotor and State 7 NABBs are robust across different HMM inferences and number of states (SI 11). Similarly, significant correlations between State 7 NABBs fractional occupancies and lifetimes and symptom severity scores were robust across all these models (SI 12).

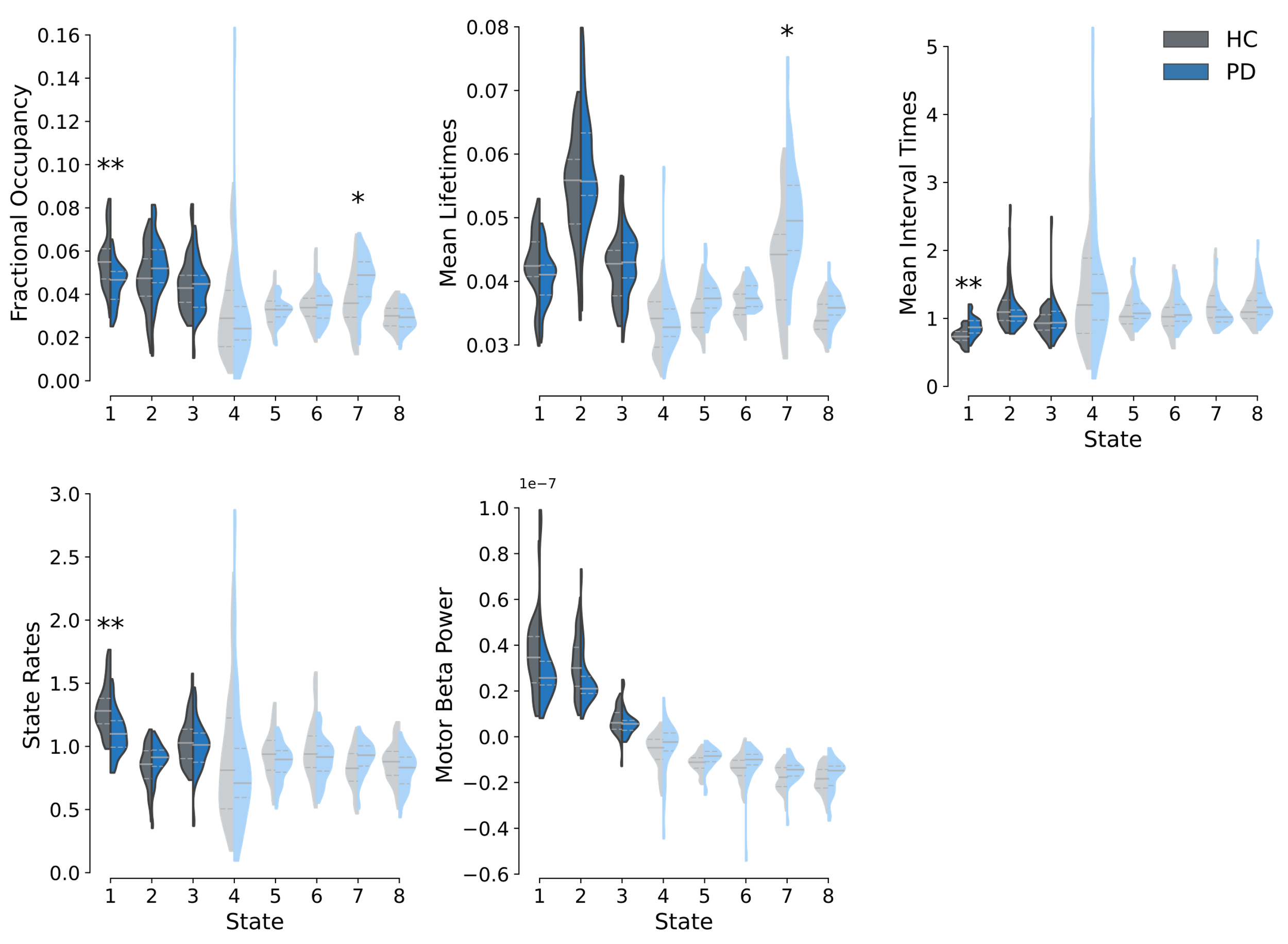

**Figure SI 9 Group comparison of network associated beta burst dynamics (NABBs) between HCs and PD patients.** Fractional occupancies, mean lifetimes, mean interval times, state rates, and motor beta power values of HCs (grey) and PD patients (blue) are depicted. Significance of group differences is assessed with maximum t-statistic permutation tests controlling for multiple comparisons across states. T-statistics were calculated from GLMs accounting for confounds*.* State metrics of NABBs of large-scale networks that have decreased motor cortical beta power are plotted in shaded colours.
** indicate p < .01 and * p < .05.

#### Different NAABs may represent different functionality

To check whether the extraction of NABBs may have the potential to disentangle beta events with different functionalities, we assessed associations between NABB fractional occupancies and other variables such as Bradykinesia/Rigidity scores and motor cortical beta power and coherence. Differing association patterns may provide evidence for a dissociation of the NABB functionality.

The analyses found significant associations between the sensorimotor NABBs’ fractional occupancies and beta power, but not beta coherence and Bradykinesia/Rigidity scores. On the other hand, no significant relationships between State 7 NABBs’ fractional occupancies and beta power were found, whereas motor cortical beta coherence and Bradykinesia/Rigidity scores were significantly linked to State 7 NAAB fractional occupancies (see SI 10).

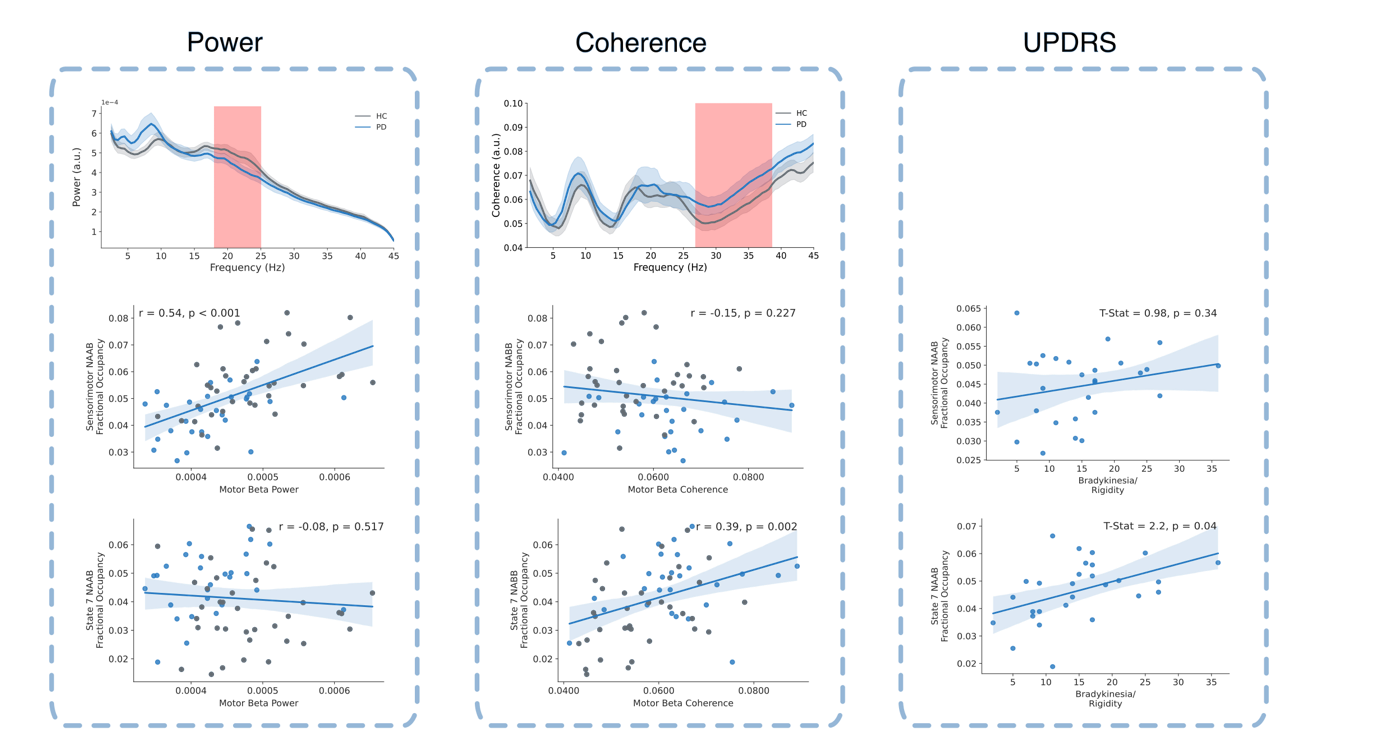

**Figure SI 10 Sensorimotor and State 7 network associated beta bursts show differing patterns of association with beta power, beta coherence, and Bradykinesia/Rigidity scores.**

These differing association patterns hint at different functionalities of the State 7 and sensorimotor NAABs, with changes in the State 7 NAAB relevant to motor symptom severity in PD. The wideband power profiles of State 7 indicate a brain wide decrease in 2 to 20Hz Power from the mean power when State 7 occurs. Since this is a somewhat unexpected finding and is difficult to explain in the context of the existing literature, these findings should be treated carefully and further replication in other studies is required.

#### Robustness of NABB Metric group contrast

Robustness checks of group contrasts of metrics of the State 7 state and sensorimotor NABBs demonstrated that the above reported significant differences are robust across all HMM fits (see SI 11).

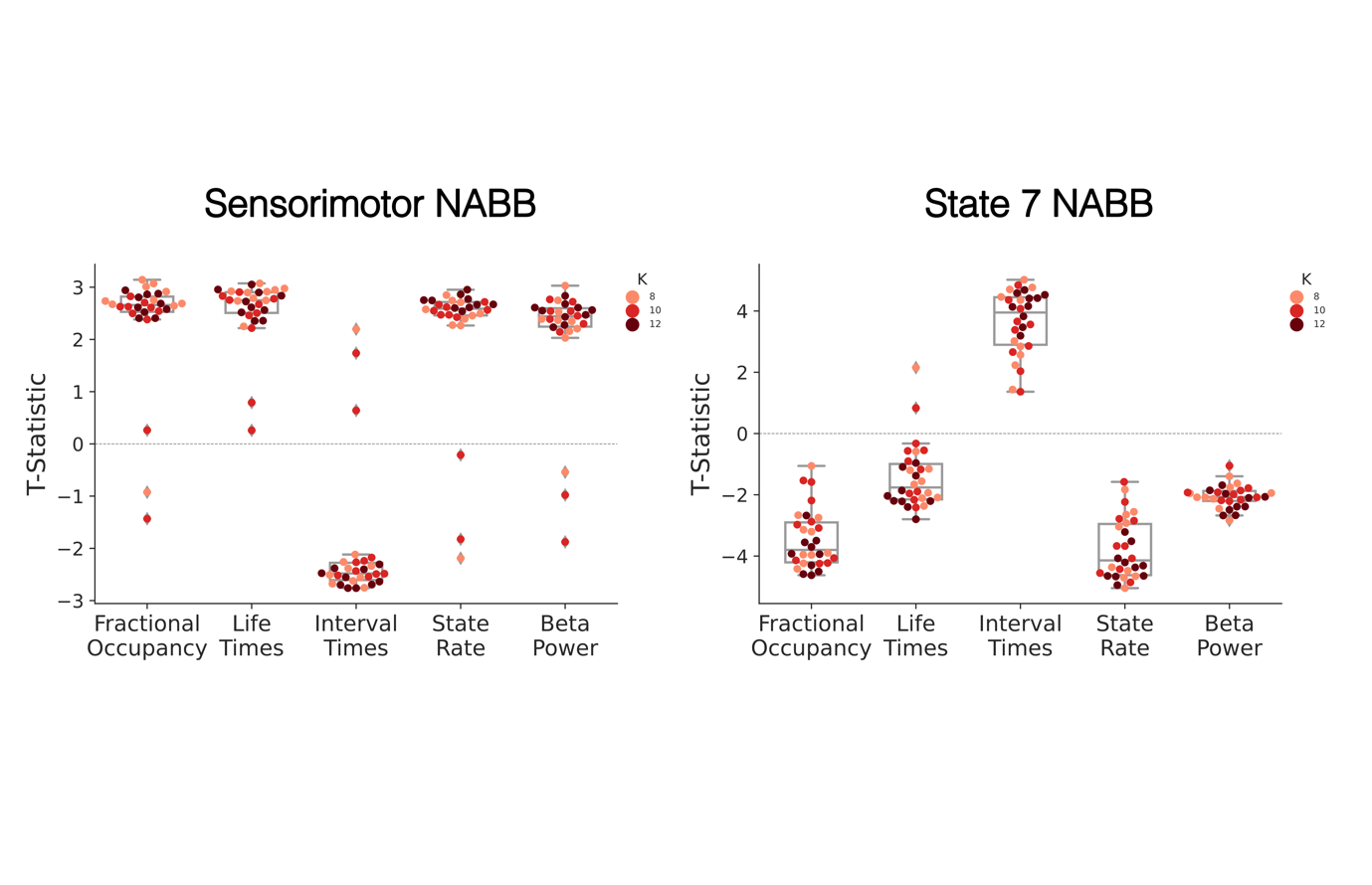

**Figure SI 11 Group differences of sensorimotor and State 7 network informed burst metrics are robust across all HMM inferences.** Each dot represents the t-statistic from the respective state metric’s group contrast between HCs and PD patients for a different HMM inference. The colours of dots indicate how many states were inferred for the depicted HMM inference.

#### Robustness of NABB Metric x UPDRS associations

Robustness checks of the association between Bradykinesia/Rigidity scores and metrics of the State 7 and sensorimotor NABBs demonstrate that associations between Bradykinesia/Rigidity scores and State 7 NABBs’ fractional occupancies and lifetimes are robust across HMM fits. No significant associations between sensorimotor NABBs and Bradykinesia/Rigidity scores were found across all HMM fits (see SI 12).

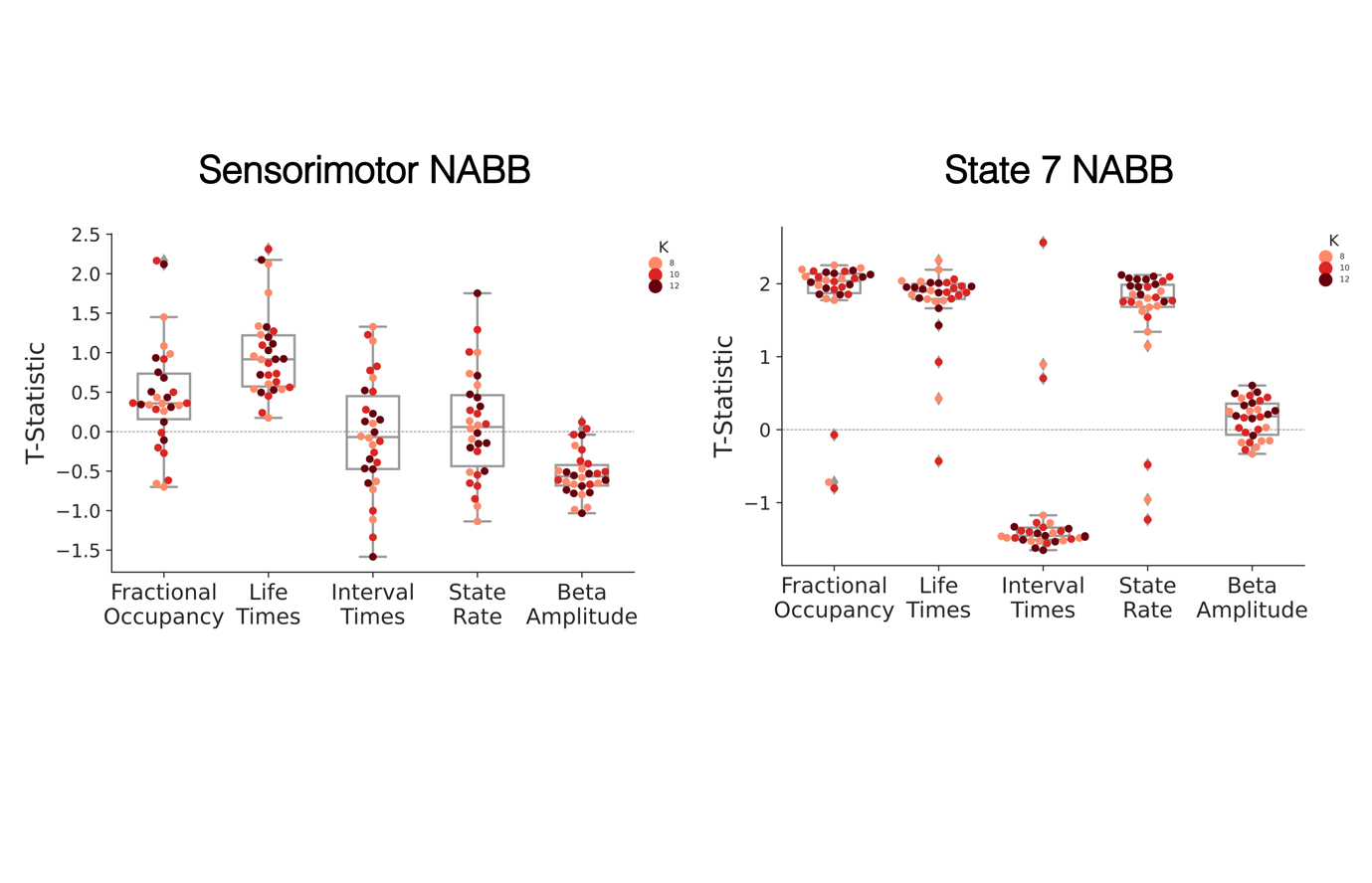

**Figure SI 12 Associations between State 7 network associated beta burst metrics and Bradykinesia/Rigidity scores is robust across all HMM inferences.** Each dot represents the t-statistic from the respective state metric’s correlation with Bradykinesia/Rigidity score for a different HMM inference. The colours of dots indicate how many states were inferred for the depicted HMM inference.
